# Rare copy number variation in alcohol use disorder using whole-genome sequencing in the All of Us Research Program

**DOI:** 10.64898/2026.11.18.26367879

**Authors:** Saeed Farajzadeh Valilou, Sydney Kramer, Masoud Edizadeh, MadhurBain Singh, Roseann E. Peterson, Christopher Chatzinakos, Silviu Alin Bacanu, Bradley T. Webb, Kenneth Kendler, Tan-Hoang Nguyen, Brien P. Riley

**Affiliations:** Virginia Institute for Psychiatric and Behavioral Genetics, Department of Psychiatry, Virginia Commonwealth University, Richmond, VA, USA; Department of Cellular, Molecular, and Genetic Medicine, Virginia Commonwealth University, Richmond, VA, USA; Social, Genetic and Developmental Psychiatry Centre, Institute of Psychiatry, Psychology and Neuroscience, King’s College London, London, UK; Department of Psychiatry and Behavioral Sciences, State University of New York System [SUNY] Downstate University, Brooklyn, NY, USA; Department of Population Health Sciences, Geisinger, Danville, PA, USA; Department of Psychiatry, Virginia Commonwealth University, Richmond, VA, USA

**Author notes:** Co-first authors (Saeed Farajzadeh Valilou, Sydney Kramer). Co-senior authors (Tan-Hoang Nguyen, Brien P. Riley).

## Abstract

Alcohol use disorder (AUD) is common, heritable (*h^2^* ∼50%) and polygenic, but the contribution of rare structural variation is largely unexamined. Using short-read whole-genome sequencing (srWGS) data from the All of Us Research Program, we tested the association of rare copy number variants (rCNV; minor allele frequency <1%) with AUD in 56,062 participants (5,410 cases, 50,652 controls) defined from electronic health records. Analyses were conducted within European-like (EUR; N=29,392), African-like (AFR; N=8,269) and Admixed American-like (AMR; N=8,049) samples and combined by cross-ancestry meta-analysis. We tested genome-wide rCNV burden, burden within AUD-related, mouse behavioral and neuropsychiatric gene sets, and individual rCNV regions. No genome-wide burden metric or individual rCNV region ≥10 kb was associated with AUD after false discovery rate (FDR) correction. Significant gene-set results were almost exclusively duplications associated with reduced AUD risk across biologically unrelated sets, suggesting a global duplication dosage effect rather than pathway-specific enrichment. In contrast, deletion count within a Neurof_UnionStringent (Neurofunction Union Stringent) gene set was associated with increased AUD risk (OR=1.04, FDR=0.044), and deletions in xenobiotic-response genes showed nominal associations in multiple ancestry groups. Among rCNV of all sizes, a 58 bp deletion at 16p12.1, a dosage-sensitive neuropsychiatric locus, was significant in the cross-ancestry analysis (OR=2.93, FDR=0.030) without significant heterogeneity. rCNV detectable by srWGS do not appear to contribute substantially to AUD liability at current sample sizes.

## Introduction

Alcohol use disorder (AUD) is a chronic condition, where individuals continue to drink excessively despite negative health and social consequences (1). AUD is a major cause of both morbidity and mortality but often goes undiagnosed and untreated (2,3). In 2021, approximately 29.5 million individuals in the United States aged 12 and older met the Diagnostic (4) and Statistical Manual of Mental Disorders, Fifth Edition (DSM-5) (1) criteria for AUD, leading to roughly 178,000 deaths (3). The lifetime prevalence of AUD is estimated at 29.1%, with nearly half of the cases classified as severe. Despite this high prevalence, <8.7% of those diagnosed received any form of treatment for AUD (5).

The genetic architecture of AUD is complex and polygenic. AUD has moderate twin-based heritability (*h^2^*) of ∼50% (6), and large-scale genome-wide association studies (GWAS) have identified associated loci (7–10). The largest GWAS of alcohol-related conditions, using a phenotype termed problematic alcohol use (PAU, including both self-report and formal diagnosis), identified more than 110 independent risk variants in 90 loci (11). Despite these successes, important gaps remain in elucidating the genetic etiology of AUD. First, even with large samples, the estimated single-nucleotide polymorphism (SNP)-based heritability (*h²_SNP_*) of AUD and PAU is relatively low (5.6% to 10.0%), indicating substantial missing heritability. Second, although samples of African ancestry are now large enough to yield significant within-ancestry signals (11,12), the majority of samples used in human genetic studies, including those focused on AUD, are derived from European ancestry. This lack of ancestral diversity limits both our understanding of the genetic basis of these traits and the generalizability of findings to global populations (8,11). Third, most associated variants are noncoding and have small effect sizes, complicating the identification of causal genes (13). Finally, AUD samples are well-powered for common variants, but lack power for rarer variation, potentially overlooking relevant genes (14).

Rare variant analyses of AUD and problematic drinking have also primarily focused on European populations. Whole-exome sequence (WES) data from the UK Biobank (UKB) identified 25 protein-coding variants and 13 genes associated with alcohol consumption (14). Gene-level analysis of the Alcohol Use Disorders Identification Test problem subscale (AUDIT-P) in the UKB WES data showed that rare variation in *ADH1C* and *THRA* is associated with AUDIT-P (15). A recent WES study identified additional genes enriched for ultra-rare variation in AUD cases (16). However, analyses of copy number variants (CNV) in AUD have previously been limited to small samples and array-based CNV (17–19). Specific rare CNV (rCNV; MAF <1%) display large effect sizes in psychiatric and neurodevelopmental disorders (20,21) including autism (22,23), schizophrenia (24), epilepsy (25), intellectual disability (26), and brain structure (27). Most prior CNV studies used SNP microarray intensity data, but the release of biobank-scale whole-genome sequence (WGS) datasets allow for large-scale WGS-based CNV studies, including of previously understudied traits like AUD, as we present here.

Phenotypic heterogeneity remains a central challenge in genetic studies of AUD. As in other complex psychiatric traits (28), phenotypic definition heavily influences genetic discoveries (8). While most AUD GWAS participants were formally ascertained cases and controls based on DSM or International Classification of Diseases (ICD) criteria, the moderate *h²_SNP_* and limited number of associated genomic loci motivate the inclusion of population biobank samples to increase discovery power. For example, quantitative assessments such as the AUDIT are available for a subset of UKB participants, and GWAS of intake/frequency (AUDIT-C) and negative consequences (AUDIT-P) yield distinct genetic architectures (9,29,30). The phenotypic challenge is particularly evident in defining AUD using electronic health records (EHR), where formal diagnoses of AUD may be absent, but other EHR data (like prescription data) suggest an individual is affected, potentially leading to misclassification (31).

Here, we used EHR-based phenotyping and WGS data from the All of Us Research Program (AoU) V8 data release to investigate rCNV associated with AUD. We first performed whole-genome burden analyses, followed by gene-set burden analyses using genes implicated in 1) AUD (11), 2) behavioral phenotypes in mouse models (32–34) and 3) curated gene-sets from a study of CNV in post-traumatic stress disorder (PTSD) (35). We estimated associations of rCNV burden and individual rCNV with AUD within three genetic ancestry groups (European-like [EUR], African-like [AFR], and Admixed American-like [AMR]) followed by cross-ancestry meta-analyses of both burden and region-level summary statistics. We also performed rCNV meta-analyses of both burden and region-level summary statistics across ancestries to increase statistical power to detect significant biological pathways associated with AUD. Additionally, we performed an exploratory analysis using trans-ancestry assignments (36) (Figure 1, Supplementary Figure 1). To our knowledge, this is the first large-scale, WGS-based rCNV analysis of AUD conducted across multiple ancestries.

**Figure 1.**
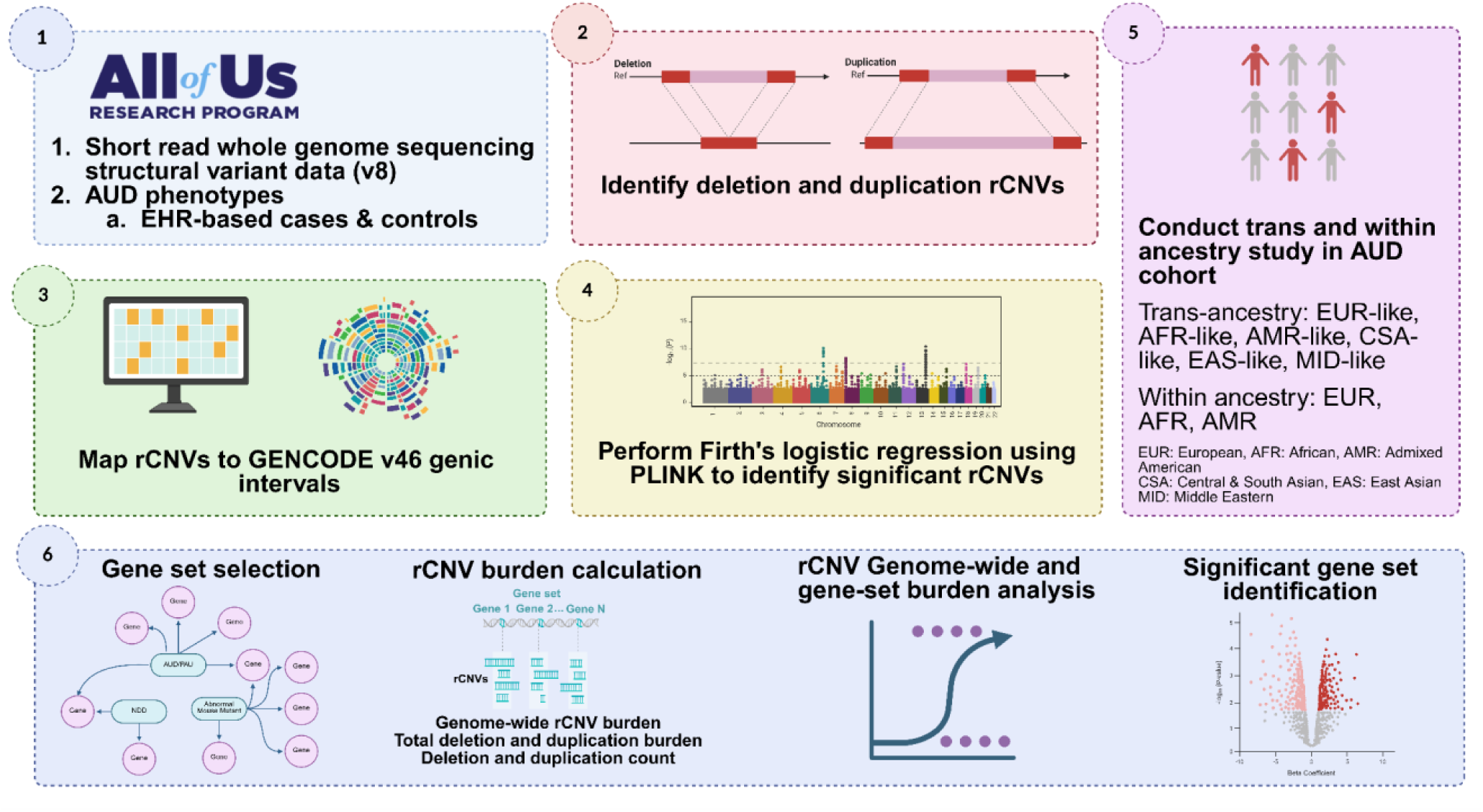
Overview of the study design and analytical workflow. All of Us srWGS structural variant data and EHR-based AUD phenotypes were used to identify deletion and duplication rCNV. rCNV were mapped to GENCODE v46 genes and tested for AUD association using Firth’s logistic regression. Trans-and within-ancestry analyses and rCNV burden analyses were performed to identify significant rCNV and gene sets.

## Data and Methods

### Participants and phenotyping

AUD cases were defined by the International Classification of Diseases, 9th/10th Revision, Clinical Modification (ICD-9/10-CM) diagnostic codes for AUD and chronic alcohol-related conditions (Supplementary Table 1), or by prescriptions for acamprosate or disulfiram in the absence of such codes (n = 136) (37,38). Controls had no AUD-related diagnoses or prescriptions and reported at least one lifetime alcoholic drink in the AoUs survey, thereby excluding individuals who reported lifetime abstinence. Individuals prescribed naltrexone without an AUD diagnosis (n = 2,695) were excluded to avoid confounding with opioid use disorder (39). After phenotypic and genomic quality control (QC), 22,385 cases and 347,195 controls remained. Of these, 56,062 (5410 cases and 50,652 controls) had CNV calls passing CNV-specific quality-control (Supplementary Methods for details about phenotyping, CNV curation and detection, Supplementary Figure 2).

### Genetic Ancestry Assignment

We reassigned AoU participants to continental ancestry groups using the population grouping by Mahalanobis distance (POP-MaD) approach (36,40) with the same reference of merged 1000 Genomes Project and Human Genome Diversity Project data (36,41); assignment, outlier pruning, and computation of principal components (PCs) are described in detail in the Supplementary Methods. Because only a subset of AoU participants have CNV calls, the East Asian-like (EAS), Central and South Asian-like (CSA), Middle Eastern-like (MID), and Oceania-like (OCE) groups each had fewer than 1000 individuals, so within-ancestry analyses included only the EUR, AFR and AMR groups (Table 1) (Supplementary Figure 3, Supplementary Table 2).

**Table 1.**
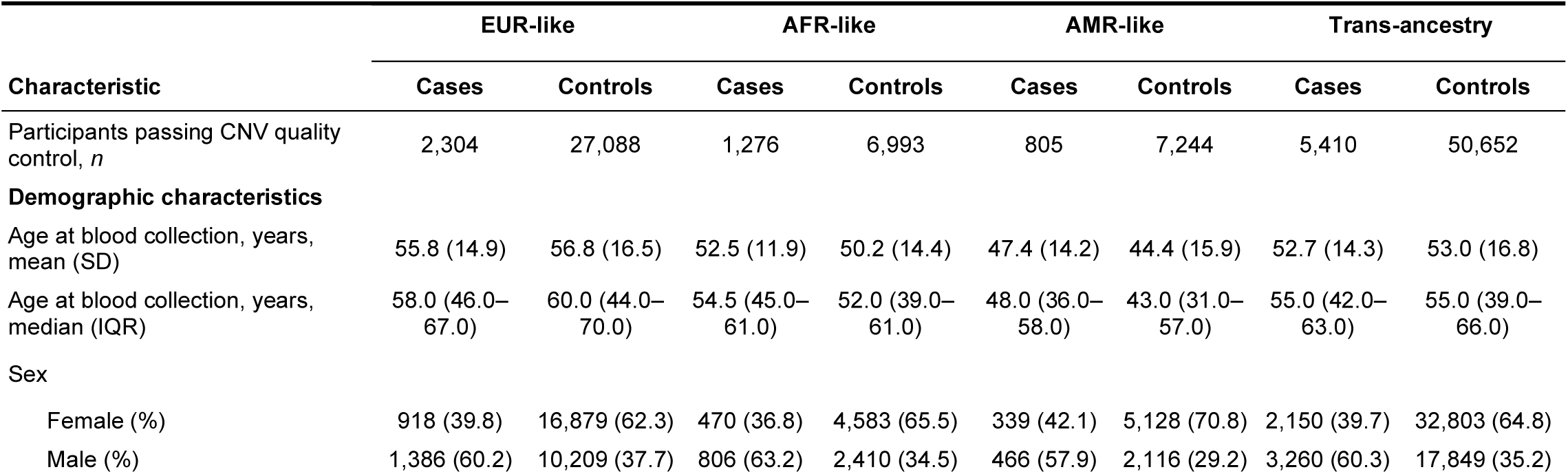
Demographics of AUD cohort analyzed in this study.

| Characteristic | **EUR-like** |  | **AFR-like** |  | **AMR-like** |  | **Trans-ancestry** |  |
|---|---|---|---|---|---|---|---|---|
|  | **Cases** | **Controls** | **Cases** | **Controls** | **Cases** | **Controls** | **Cases** | **Controls** |
| Participants passing CNV quality control, *n* | 2,304 | 27,088 | 1,276 | 6,993 | 805 | 7,244 | 5,410 | 50,652 |
| **Demographic characteristics** |  |  |  |  |  |  |  |  |
| Age at blood collection, years, mean (SD) | 55.8 (14.9) | 56.8 (16.5) | 52.5 (11.9) | 50.2 (14.4) | 47.4 (14.2) | 44.4 (15.9) | 52.7 (14.3) | 53.0 (16.8) |
| Age at blood collection, years, median (IQR) | 58.0 (46.0–67.0) | 60.0 (44.0–70.0) | 54.5 (45.0–61.0) | 52.0 (39.0–61.0) | 48.0 (36.0–58.0) | 43.0 (31.0–57.0) | 55.0 (42.0–63.0) | 55.0 (39.0–66.0) |
| Sex |  |  |  |  |  |  |  |  |
| Female (%) | 918 (39.8) | 16,879 (62.3) | 470 (36.8) | 4,583 (65.5) | 339 (42.1) | 5,128 (70.8) | 2,150 (39.7) | 32,803 (64.8) |
| Male (%) | 1,386 (60.2) | 10,209 (37.7) | 806 (63.2) | 2,410 (34.5) | 466 (57.9) | 2,116 (29.2) | 3,260 (60.3) | 17,849 (35.2) |

### CNV burden analysis

CNV burden was assessed at both the genome-wide and gene-set levels, following a previously published analytic framework (35). CNV burden was quantified using multiple complementary metrics, including: (1) the cumulative length in megabases (Mb) of all rare duplications and deletions analyzed separately; (2) the number of rare duplications and deletions considered separately; and (3) the aggregate length of all duplications and deletions combined, and evaluated for association with AUD using Firth’s logistic regression. We initially analyzed CNV ≥10 Kb due to their greater potential for pathogenicity, and then rCNV of any size. We conducted genome-wide and gene-set burden analyses within the three largest ancestry groups (EUR, AFR, AMR), meta-analyzed the within-ancestry results, and performed a trans-ancestry analysis of all samples pooled together (rationale in Supplementary Methods).

#### Genome-wide burden analysis

We conducted whole-genome rCNV burden analyses for both length and count of deletions and duplications separately, and cumulative length of all rCNV metrics using Firth’s logistic regression, adjusted for the first ten genetic principal components to control for population stratification. We tested three covariate models for genome-wide burden analyses: 1) the first ten PCs only; 2) the first ten PCs, age (at blood draw), sex; and 3) the first ten PCs, age, sex, total rCNV count, and average rCNV length (Supplementary Figure 4). Effect estimates were broadly consistent across models. Following previous studies (35,42) we report Model 3 as the primary model in the main text because it is the most conservative, additionally accounting for individual differences in global rCNV load.

#### Gene-set burden analysis

At the gene-set level, analyses quantified the extent to which rCNV overlapped individual genes and predefined gene sets. To map genes to each identified CNV to calculate burden in Mb, we used the GENCODE Release 46 (GRCh38.p14) (43) genic intervals (Supplementary Table 10). Each CNV was assigned to any genes sharing at least one base pair of overlap (35).

Gene-set burden analysis included 5 AUD/PAU related gene sets from a previous PAU study (11) plus a combination of all five sets, 146 abnormal behavior gene sets derived from mouse mutants with central nervous system (CNS) phenotypes (32–34), 73 curated sets tested in the PGC PTSD study and one negative control cholesterol gene-set, previously shown to have no genetic association with PAU (11) (Supplementary Table 11). Each CNV was assigned to all genes with which it shared at least one base pair of overlap. For each gene, the total number of overlapping CNV and the extent of base pair overlap were calculated (Supplementary Table 10). Gene burden was encoded as a binary variable, where individuals were assigned a value of 1 if at least one CNV overlapped the gene and 0 otherwise.

We adjusted for average rCNV length and total rCNV count to reduce confounding by background genomic instability. Total rCNV count accounts for differences in the rate of rCNV formation between cases and controls, whereas average rCNV length accounts for differences in the size of the events themselves (42) (Supplementary Methods). Our model followed previously established frameworks (35) and tested five gene-set level burden metrics: deletion/duplication count, deletion/duplication length, and total length within each gene set. For each metric, we fitted Firth’s logistic regression model of the following form, adjusting for genome-wide average rCNV length, genome-wide total rCNV count, the first ten within-ancestry genetic PCs, age, and sex.

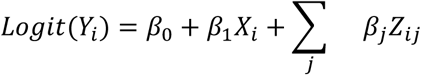

where *Y_i_* represents the case/control status of individual *i* (1 = case, 0 = control). *β*_0_ represents the intercept. *β*_1_ represents the coefficient of *X_i_*. *X_i_* represents the count or length of rCNV. *β_j_* represents the corresponding coefficient for covariate j and *Z_ij_* represents the value for the covariate j for individual *i*.

#### Cross-ancestry burden meta-analysis

Ancestry-specific *β* estimates and their standard errors (SEs) were combined across ancestries (EUR, AFR and AMR) using fixed-effects meta-analysis under two complementary frameworks. First, a sample-size– weighted Stouffer’s Z (44) was computed because it weights each ancestry by the square root of its sample size 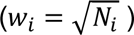, which is robust to imbalance in cohort size, and an inverse-variance–weighted (IVW) fixed-effects estimate, which provides pooled odds ratios (OR) and 95% confidence intervals (CIs). Formulas are provided in the Supplementary Methods.

#### Individual rCNV analysis

Individual rCNV associations with AUD were evaluated using Firth’s logistic regression to account for sparse data and low-frequency events. Primary ancestry-stratified analyses were conducted within the EUR, AFR, and AMR groups. We also performed an exploratory analysis in a trans-ancestry framework using all QC-passed samples from all ancestry groups (adding CSA, EAS, and MID). For each model, age, sex, and the first 10 within-ancestry PCs (EUR, AFR and AMR) or trans-ancestry PCs were covariates. We analyzed CNV regions at two size thresholds: ≥10 kb and all CNV sizes.

#### Individual rCNV meta-analysis

Association results for individual CNV regions in the EUR, AFR, and AMR ancestry groups were meta-analyzed using METAL (45) with the sample-size weighted scheme. To account for imbalanced case-control ratios, the effective sample size (*N_eff_*; Supplementary Methods) was used as the sample size for each study. Genomic control correction was applied. Between-ancestry heterogeneity was assessed using Cochran’s *Q* and *I*². Significance was assessed after FDR correction across all regions tested within CNV size thresholds (all CNV sizes and ≥10kb CNV).

## Results

We analyzed rCNV called from AoU srWGS data on 56,062 participants passing CNV QC, comprising 5,410 AUD cases and 50,652 controls. Ancestry-stratified analyses were conducted in the three largest groups, EUR (N = 29,392; 2,304 cases and 27,088 controls), AFR (N = 8,269; 1,276 cases and 6,993 controls) and AMR (N = 8,049; 805 cases and 7,244 controls), together with a trans-ancestry analysis of all pooled samples (N = 56,062; 5,410 AUD cases and 50,652 controls). Because of their greater potential for pathogenicity and because short calls are the most sensitive to breakpoint uncertainty in short-read data, rCNV of at least 10 kb were treated as the primary analysis set, and all CNV sizes were analyzed secondarily. We evaluated rCNV at five levels of resolution: 1) genome-wide rCNV burden, and burden within 2) six curated AUD and PAU gene sets, 3) 146 abnormal mouse behavior gene sets, and 4) 73 curated gene sets from a previous PGC PTSD study of CNV; and 5) association of individual rCNV regions with AUD.

### Genome-wide rCNV burden

First we asked whether the total genome-wide load of rCNV differed between cases and controls, using five burden metrics (deletion, duplication and total burden in Mb, and deletion and duplication counts) adjusted for the first ten within-ancestry principal components. For rCNV of ≥10 kb, no metric was associated with AUD in any ancestry group or in the trans-ancestry analysis (all FDR > 0.40), and the cross-ancestry meta-analysis was also null (Supplementary Tables 3 and 4). When CNV of all sizes were included, deletion count was associated with AUD in the EUR sample (OR = 1.004, 95% CI 1.003-1.005, *P* = 9.7 × 10^-9^, FDR = 4.9 × 10^-8^) and in the trans-ancestry analysis (OR = 1.0014, 95% CI 1.0007-1.0021, *P* = 4.4 × 10^-5^, FDR = 2.2 × 10^-4^), with no corresponding signal in the AFR (*P* = 0.90) or AMR (*P* = 0.54) samples. The meta-analysis of deletion count across the three ancestries was significant (Stouffer Z = 4.90, *P_meta_*= 9.4 × 10^-7^, FDR = 0.002) but highly heterogeneous (*I^2^* = 90.1%, *P* heterogeneity < 0.001), indicating that the result was driven by the EUR sample. Because this signal was confined to the unrestricted size category, in which short calls predominate, and disappeared when the analysis was limited to rCNV ≥10 kb, we regard it as most consistent with ancestry-specific differences in the detection of short deletions rather than with a genuine excess of genome-wide deletion load in cases (Supplementary Table 5). In the EUR-like sample, duplication count for rCNV of all sizes was nominally significant before, but not after, multiple-testing correction (OR = 1.0011, 95% CI 1.0002 to 1.0021, *P* = 2.3 × 10^-2^, FDR = 5.7 × 10^-2^). We did not find any significant burden length variables in any test (Table 2).

**Table 2.** Genome-wide burden results.

| Ancestry | Size Filter | Burden Metric | β | SE | Z | OR (95% CI) | *P* | FDR *P* |
|---|---|---|---|---|---|---|---|---|
| EUR-like | All sizes | Deletion Count | 0.004 | 0.0007 | 5.7274 | 1.004 (1.0026-1.0054) | 9.73E-09 | **4.86E-08** |
|  |  | Duplication Count | 0.0011 | 0.0005 | 2.2716 | 1.0011 (1.0002-1.0021) | 2.29E-02 | 5.73E-02 |
| AFR-like | ≥10kb | Duplication Burden (Mb) | 0.0213 | 0.0174 | 1.2258 | 1.0216 (0.9873-1.057) | 1.99E-01 | 6.20E-01 |
| Trans-ancestry | ≥10kb | Deletion Burden (Mb) | 0.0174 | 0.0127 | 1.3673 | 1.0176 (0.9925-1.0433) | 1.63E-01 | 4.07E-01 |
|  |  | Total Burden (Mb) | 0.0096 | 0.0068 | 1.3993 | 1.0096 (0.9962-1.0233) | 1.60E-01 | 4.07E-01 |
|  | All sizes | Deletion Burden (Mb) | 0.0184 | 0.0124 | 1.4777 | 1.0185 (0.994-1.0436) | 1.32E-01 | 1.77E-01 |
|  |  | Total Burden (Mb) | 0.0098 | 0.0067 | 1.4639 | 1.0098 (0.9967-1.0232) | 1.42E-01 | 1.77E-01 |
|  |  | Deletion Count | 0.0014 | 0.0004 | 4.0844 | 1.0014 (1.0007-1.0021) | 4.36E-05 | 2.18E-04 |
|  |  | Duplication Count | 0.0006 | 0.0003 | 1.8593 | 1.0006 (1-1.0012) | 6.28E-02 | 1.57E-01 |

### Gene-set burden analysis

Genome-wide total rCNV count and average rCNV length were carried forward as covariates in all subsequent gene-set analyses, following previous studies (35,42).

#### Burden within AUD and PAU gene sets

We then tested the six gene sets derived from the most recent large-scale PAU study (11), comprising the MAGMA, fine-mapped TWAS, H-MAGMA chromatin interaction, SMultiXcan and SPrediXcan sets and a combined set uniting all five. No burden metric reached FDR significance in any ancestry group or CNV size category. Among rCNV ≥10 kb, the strongest result was an association between duplication count in the SMultiXcan set and AUD in the trans-ancestry analysis (OR = 0.87, 95% CI 0.76-0.99, *P* = 0.038, FDR = 0.22). For CNV of all sizes, deletion burden was elevated in the trans-ancestry analysis both for the combined set (OR = 1.05, 95% CI 1.01-1.10, *P* = 0.010, FDR = 0.099) and for the H-MAGMA chromatin interaction set defined across six brain annotations (OR = 1.05, 95% CI 1.01-1.10, *P* = 9.9 × 10^-3^, FDR = 0.099), and deletion burden was positive in direction for every individual AUD set in the EUR-like sample (*P* between 0.020 and 0.029) (Supplementary Tables 6 and 7). Cross-ancestry meta-analysis of these sets produced no result surviving correction under either the Stouffer or the inverse-variance weighted framework (Supplementary Table 8 and 9).

#### Burden across abnormal mouse behavior and curated gene sets from PGC PTSD CNV study

Each of the gene sets was tested with five burden metrics per ancestry group and CNV size category. For rCNV of at least 10 kb in size, no test reached FDR significance in the EUR, AFR, or AMR samples analyzed separately. The strongest results in the EUR sample were inverse associations with duplication metrics in the abnormal mouse behavior sets: human postsynaptic density core set (burden OR = 0.60, 95% CI 0.47-0.77, *P* = 7.4 × 10^-5^, FDR = 0.057; count OR = 0.89, 95% CI 0.83 to 0.94, P = 1.5 × 10^-4^, FDR = 0.057), the postsynaptic density Bayesian set (count OR = 0.93, 95% CI 0.90-0.97, *P* = 1.4 × 10^-4^, FDR = 0.057) and the abnormal nervous system development set (burden OR = 0.65, 95% CI 0.51-0.81, *P* = 2.1 × 10^-4^, FDR = 0.061) (Table 3). No test approached significance in the AFR or AMR samples, where the smallest FDR values were 0.9999 and 0.998, respectively.

**Table 3.** Gene-set burden analysis results in EUR ancestry group.

| Size Filter | Burden Metric | Gene Set | β | SE | Z | OR (95% CI) | *P* | FDR *P* |
|---|---|---|---|---|---|---|---|---|
| ≥10kb | Duplication Burden (Mb) | PSD_human_core | -0.5078 | 0.1285 | -3.9504 | 0.6018 (0.4678-0.7743) | 7.43E-05 | 5.66E-02 |
|  |  | abnormal_nervous_system_development | -0.4356 | 0.1178 | -3.6968 | 0.6469 (0.5135-0.8149) | 2.09E-04 | 6.08E-02 |
|  | Duplication Count | PSD_human_core | -0.1191 | 0.0314 | -3.7899 | 0.8877 (0.8347-0.9441) | 1.46E-04 | 5.66E-02 |
|  |  | PSD_BayesGrant_fullset | -0.0729 | 0.0192 | -3.7955 | 0.9297 (0.8953-0.9654 | 1.43E-04 | 5.66E-02 |
|  | Deletion Count | abnormal_behavioral_response_to_xenobiotic | 0.277 | 0.099 | 2.7979 | 1.3192 (1.0865-1.6018 | 4.95E-03 | 9.45E-02 |
| All sizes | Duplication Burden (Mb) | PSD_human_core | -0.5029 | 0.1294 | -3.8869 | 0.6048 (0.4693-0.7794 | 9.77E-05 | 2.41E-02 |
|  |  | abnormal_nervous_system_development | -0.4334 | 0.1182 | -3.6659 | 0.6483 (0.5142-0.8174 | 2.38E-04 | 3.77E-02 |
|  | Duplication Count | abnormal_behavior | -0.0235 | 0.0064 | -3.6473 | 0.9768 (0.9645-0.9892 | 2.57E-04 | 3.77E-02 |
|  |  | BspanHM_lg2rpkm3.21 | -0.0141 | 0.0038 | -3.6812 | 0.986 (0.9786-0.9934 | 2.25E-04 | 3.77E-02 |
|  |  | BspanVHM_PstNat | -0.0185 | 0.0048 | -3.8844 | 0.9817 (0.9726-0.9909 | 9.90E-05 | 2.41E-02 |
|  |  | BspanVH_lg2rpkm4.74 | -0.0161 | 0.0042 | -3.8743 | 0.984 (0.9761-0.9921 | 1.03E-04 | 2.41E-02 |
|  |  | GOP_lessthan500g | -0.0075 | 0.0019 | -3.9083 | 0.9926 (0.9889-0.9963 | 8.99E-05 | 2.41E-02 |
|  |  | blueModule | -0.0192 | 0.0049 | -3.9014 | 0.981 (0.9716-0.9905) | 9.27E-05 | 2.41E-02 |

Gene-set definitions: PSD, postsynaptic density; PSD and (abnormal\_\*) sets comprise human orthologs of genes annotated to the corresponding Mammalian Phenotype Ontology term sets are derived from Abnormal Mouse behavior phenotype sets. Bspan sets are defined by BrainSpan brain expression level (VH, very high; HM, high–medium; VHM, very high to medium; thresholds in log2 RPKM) or developmental bias (PstNat, postnatal). See supplementary information.

In the trans-ancestry analysis, which carries approximately twice the case count of the EUR-like sample, 91 of 1,165 tests were significant after FDR correction at the 10 kb threshold, distributed across 74 gene sets (Supplementary Tables 6 and 7). The signal was largely driven by duplication metrics with uniformly betas uniformly <0 (inverse in direction), with duplication count significant in 69 sets and duplication burden in 21.

The strongest associations were with abnormal nervous system development (burden OR = 0.75, 95% CI 0.65 to 0.86, *P* = 2.8 × 10^-5^, FDR = 0.012; count OR = 0.93, 95% CI 0.90-0.96, *P* = 3.1 × 10^-5^, FDR = 0.012), genes with high-to-medium Brain-Span expression (BspanHM_lg2rpkm3.21: Genes with at least 5 BrainSpan data points log2 (rpkm) >= 3.21, but not in BspanVH_lg2rpkm4.74) (count OR = 0.98, 95% CI 0.97 to 0.99, *P* = 2.9 × 10^-5^, FDR = 0.012), abnormal associative learning (count OR = 0.83, 95% CI 0.76 to 0.91, *P* = 1.3 × 10^-4^, FDR = 0.017) and abnormal social or conspecific interaction (count OR = 0.88, 95% CI 0.83 to 0.94, *P* = 7.0 × 10^-5^, FDR = 0.017). Only one deletion metric passed correction: deletion count within the stringent neurofunction union set was associated with increased odds of AUD (OR = 1.04, 95% CI 1.01 to 1.06, *P* = 2.9 × 10^-3^, FDR = 0.044) (Supplementary Figure 5).

The cholesterol gene set, included as a negative control because cholesterol traits show no genetic correlation with problematic alcohol use (11), was not associated with AUD for any burden metric in any ancestry group or CNV size groups (minimum FDR = 0.55). For deletions it was null (trans-ancestry deletion burden OR = 1.00, *P* = 0.93 for rCNVs ≥10 kb). For duplications it was also non-significant, although its estimates lay in the same inverse direction and within the range of the sets that reached significance (duplication count OR = 0.89, *P* = 0.16); however, at 104 genes, it is an order of magnitude smaller than the largest sets tested.

The deletion results followed a narrower and more interpretable pattern. Deletion count within the abnormal behavioural response to xenobiotic set, a category that captures organismal responses to foreign chemical compounds including alcohol, was nominally associated with increased odds of AUD in the EUR sample (OR = 1.32, 95% CI 1.09 to 1.60, P = 4.9 × 10^-3^, FDR = 0.095), the AMR sample (OR = 1.46, 95% CI 1.10 to 1.95, P = 8.5 × 10^-3^, FDR = 0.998) and the trans-ancestry analysis (OR = 1.16, 95% CI 1.03 to 1.30, P = 0.015, FDR = 0.113) (Table 3, Supplementary Table 6).

### Cross-ancestry burden meta-analysis

Combining EUR-like, AFR-like, and AMR-like estimates by sample-size-weighted Stouffer meta-analysis identified eight gene-set tests surviving FDR correction at the 10 kb threshold, all of them duplication metrics and all with betas <0 (Table 4, Supplementary Tables 8 and 9). These comprised duplication burden and duplication count in abnormal nervous system development (OR = 0.83, 95% CI 0.72 to 0.95, *P* = 7.7 × 10^-5^, FDR = 0.045; OR = 0.93, 95% CI 0.90 to 0.97, *P* = 2.5 × 10^-4^, FDR = 0.045) and duplication count in abnormal associative learning (OR = 0.82, 95% CI 0.74 to 0.91, *P* = 9.0 × 10^-5^), abnormal telencephalon morphology (OR = 0.88, 95% CI 0.83 to 0.94, *P* = 1.6 × 10^-4^), abnormal cerebrum morphology (OR = 0.86, 95% CI 0.79 to 0.93, *P* = 2.6 × 10^-4^), abnormal social or conspecific interaction (OR = 0.89, 95% CI 0.83 to 0.95, *P* = 2.9 × 10^-4^), abnormal response to novelty (OR = 0.82, 95% CI 0.73 to 0.92, *P* = 2.9 × 10^-4^) and abnormal temporal lobe morphology (OR = 0.84, 95% CI 0.76 to 0.93, *P* = 4.0 × 10^-4^). Seven of the eight showed a consistent direction of effect in all three ancestries with no evidence of heterogeneity; duplication burden in abnormal nervous system development was the exception (I^2^ = 84.5%, P heterogeneity = 0.002). None of these results survived correction under the inverse-variance weighted framework, in which the smallest adjusted *P* value at this threshold was 0.084. For rCNVs of all sizes, three tests were also FDR significant under the inverse-variance weighted framework, each of them duplication count within a BrainSpan expression set: BspanHM_lg2rpkm3.21 (genes with high-to-medium (HM) brain expression) (OR = 0.989, 95% CI 0.984 to 0.994, *P* = 1.8 × 10⁻⁵, FDR = 0.034), BspanVH_lg2rpkm4.74 (OR = 0.988, 95% CI 0.982 to 0.994, *P* = 4.0 × 10⁻⁵, FDR = 0.034) and BspanVHM_PreNat (genes in the very-high/high-medium (VHM) expression tier whose expression is mainly prenatal) (OR = 0.982, 95% CI 0.973 to 0.991, *P* = 4.4 × 10⁻⁵, FDR = 0.034). All three were also significant under the Stouffer framework (FDR = 0.010, 0.010 and 0.014), showed a consistent direction of effect across the three ancestry groups, and showed no evidence of between-ancestry heterogeneity (I² = 0 to 4.8%, *P* heterogeneity 0.35 to 0.70) (Supplementary Figures 6 and 7).

**Table 4.** Cross-ancestry meta-analysis.

| Size | Burden Metric | Gene Set | Z (Stouffer Meta) | P (Stouffer Meta) | Q (Stouffer Meta) | Beta (IVW Meta) | SE (IVW Meta) | P (IVW Meta) | Q (IVW Meta) | OR (95% CI) | Cochran's Q | I² (%) | P het |
|---|---|---|---|---|---|---|---|---|---|---|---|---|---|
| ≥10kb | Duplication Burden (Mb) | abnormal_nervous_system_development | -3.9542 | 7.68E-05 | 4.52E-02 | -0.1885 | 0.0688 | 6.16E-03 | 1.39E-01 | 0.8282 (0.72–0.95) | 12.877 | 84.4684 | 1.60E-03 |
|  | Duplication Count | abnormal_associative_learning | -3.9158 | 9.01E-05 | 4.52E-02 | -0.1998 | 0.0546 | 2.52E-04 | 8.37E-02 | 0.8189 (0.74–0.91) | 3.4088 | 41.328 | 1.82E-01 |
|  |  | abnormal_telencephalon_morphology | -3.7785 | 1.58E-04 | 4.52E-02 | -0.1227 | 0.0331 | 2.10E-04 | 8.37E-02 | 0.8845 (0.83–0.94) | 0.6811 | 0 | 7.11E-01 |
|  |  | abnormal_nervous_system_development | -3.667 | 2.45E-04 | 4.54E-02 | -0.0678 | 0.0194 | 4.59E-04 | 8.37E-02 | 0.9344 (0.90–0.97) | 2.3113 | 13.4692 | 3.15E-01 |
|  |  | abnormal_cerebrum_morphology | -3.653 | 2.59E-04 | 4.54E-02 | -0.1546 | 0.0444 | 4.97E-04 | 8.37E-02 | 0.8568 (0.79–0.93) | 1.3841 | 0 | 5.01E-01 |
|  |  | abnormal_socialORconspecific_interaction | -3.6261 | 2.88E-04 | 4.54E-02 | -0.1173 | 0.035 | 8.08E-04 | 9.04E-02 | 0.8893 (0.83–0.95) | 2.8183 | 29.0353 | 2.44E-01 |
|  |  | abnormal_response_to_novelty | -3.6241 | 2.90E-04 | 4.54E-02 | -0.1976 | 0.0566 | 4.81E-04 | 8.37E-02 | 0.8207 (0.73–0.92) | 1.2199 | 0 | 5.43E-01 |
|  |  | abnormal_temporal_lobe_morphology | -3.5431 | 3.96E-04 | 4.65E-02 | -0.1753 | 0.0527 | 8.73E-04 | 9.04E-02 | 0.8392 (0.76–0.93) | 1.6825 | 0 | 4.31E-01 |
| All sizes | Duplication Burden (Mb) | abnormal_nervous_system_development | -3.9777 | 6.96E-05 | 2.34E-02 | -0.1717 | 0.0649 | 8.19E-03 | 3.65E-01 | 0.8422 (0.74–0.96) | 12.9861 | 84.5989 | 1.50E-03 |
|  | Duplication Count | BspanHM_lg2rpkm3.21 | -4.2924 | 1.77E-05 | 1.04E-02 | -0.0113 | 0.0026 | 1.78E-05 | 3.43E-02 | 0.9888 (0.98–0.99) | 1.5196 | 0 | 4.68E-01 |
|  |  | BspanVHM_PreNat | -4.1694 | 3.05E-05 | 1.44E-02 | -0.0183 | 0.0045 | 4.39E-05 | 3.43E-02 | 0.9819 (0.97–0.99) | 0.7031 | 0 | 7.04E-01 |
|  |  | BspanVH_lg2rpkm4.74 | -4.2966 | 1.73E-05 | 1.04E-02 | -0.0121 | 0.0029 | 3.98E-05 | 3.43E-02 | 0.9880 (0.98–0.99) | 2.1 | 4.7641 | 3.50E-01 |
|  |  | BspanVHM_PstNat | -3.9839 | 6.78E-05 | 2.34E-02 | -0.0127 | 0.0034 | 1.59E-04 | 5.34E-02 | 0.9874 (0.98–0.99) | 2.9657 | 32.5624 | 2.27E-01 |
|  |  | GOP_lessthan500g | -3.8991 | 9.66E-05 | 2.64E-02 | -0.0051 | 0.0013 | 1.33E-04 | 5.19E-02 | 0.9949 (0.99–1.00) | 4.5035 | 55.5903 | 1.05E-01 |
|  |  | ALL_UNIQUE_146_NDD_SETS | -3.6829 | 2.31E-04 | 4.17E-02 | -0.0056 | 0.0015 | 2.37E-04 | 6.95E-02 | 0.9945 (0.99–1.00) | 2.1066 | 5.0622 | 3.49E-01 |

Gene-set definitions: Abnormal mouse behavior phenotype set: abnormal nervous system development; abnormal associative learning; abnormal telencephalon morphology; abnormal cerebrum morphology; abnormal social/conspecific interaction; abnormal response to novelty; abnormal temporal lobe morphology. Brain expression sets (BrainSpan): BspanVH\_lg2rpkm4.74, genes with very high brain expression (log2 RPKM ≥ 4.74); BspanHM\_lg2rpkm3.21, genes with high-to-medium brain expression (log2 RPKM ≥ 3.21 and < 4.74); BspanVHM\_PreNat, very-high- and high-to-medium-expressed genes with higher prenatal than postnatal expression; BspanVHM\_PstNat, very-high- and high-to-medium-expressed genes with higher postnatal than prenatal expression.

### Association of individual rCNV with AUD

No rCNV region of at least 10 kb was associated with AUD after FDR correction in the EUR, AFR or AMR samples, in the trans-ancestry analysis, or in the cross-ancestry meta-analysis. When rCNV of any size were considered, two deletions reached FDR significance in the trans-ancestry analysis. A 58 bp deletion at 16p12.1 overlapping *ENSG00000260580* gave the smallest *P* value genome-wide (OR = 2.93, 95% CI 1.94 to 4.42, P = 3.06 × 10-7, FDR = 0.030), and a 171 bp intergenic deletion at 2p11.2 was also significant (OR = 14.99, 95% CI 5.15 to 43.65, *P* = 6.90 × 10-7, FDR = 0.034). The 16p12.1 deletion was likewise the top-ranked region in the EUR sample (OR = 4.18, 95% CI 2.28 to 7.65, *P* = 3.70 × 10-6, FDR = 0.064), where three further regions approached significance: a 582 bp intergenic duplication at 2p11.2 (OR = 3.42, 95% CI 2.05 to 5.71, *P* = 2.38 × 10-6, FDR = 0.064), a 735 bp deletion at 2q37.3 overlapping *ESPNL* (OR = 3.38, 95% CI 2.03 to 5.63, *P* = 2.99 × 10-6, FDR = 0.064) and a 159 bp deletion at 22q13.31 overlapping *TBC1D22A* (OR = 7.41, 95% CI 3.13 to 17.56, *P* = 5.39 × 10-6, FDR = 0.070). No region survived FDR correction in the AFR-like or AMR-like samples, possibly consistent with their smaller case counts. In the cross-ancestry meta-analysis of the three groups, the 16p12.1 deletion again ranked first (Z = 4.81, *P* = 1.54 × 10⁻⁶, FDR = 0.10), although no region passed correction (FDR < 0.05). There was no significant between-ancestry heterogeneity at this locus (I² = 33.9%, P heterogeneity = 0.22). All implicated events were very short and uncommon, and each conferred a roughly three-fold to fifteen-fold increase in the odds of AUD among carriers (Table 5, Supplementary Figure 8).

**Table 5.**
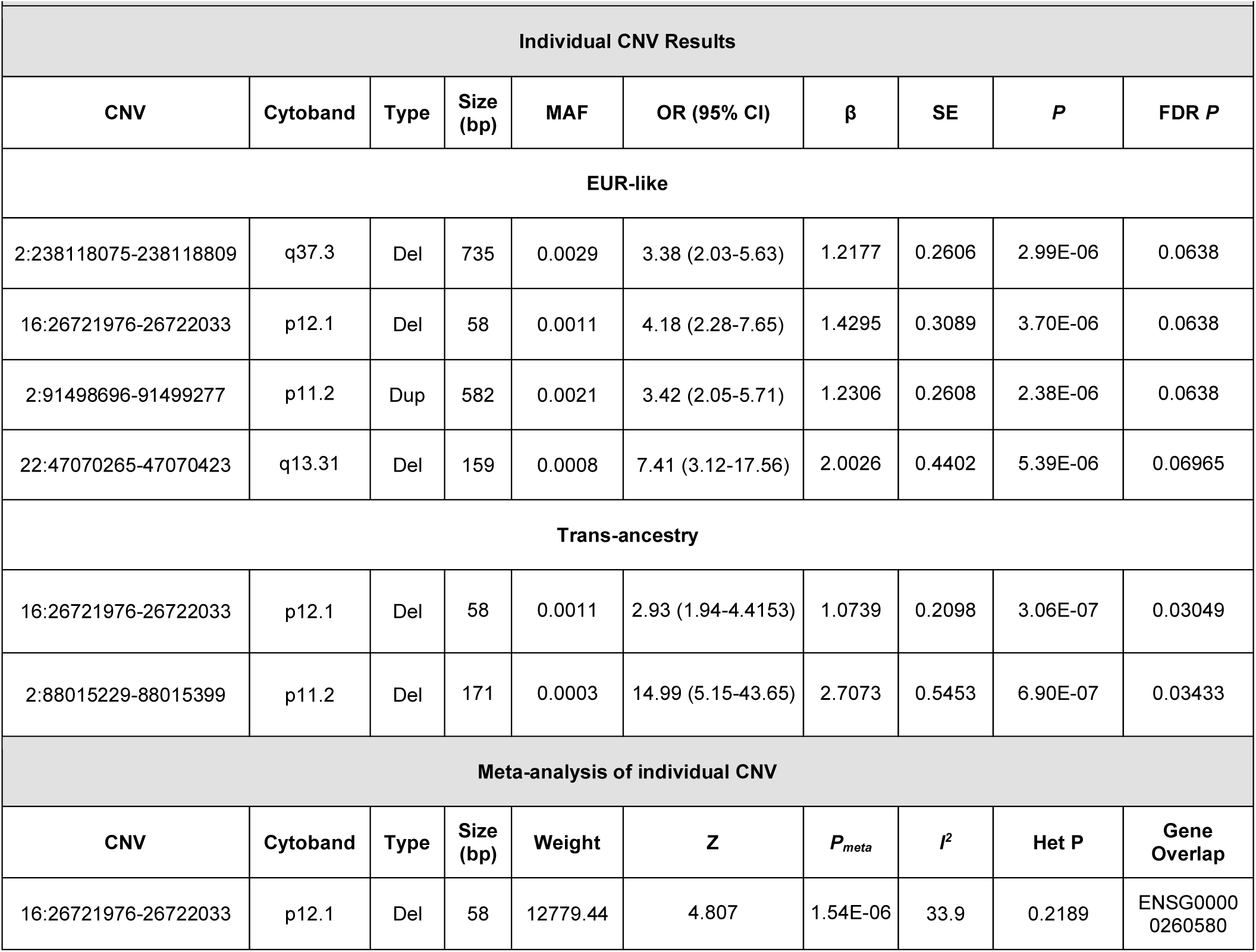
Individual CNV region and meta analysis results.

| **Individual CNV Results** |  |  |  |  |  |  |  |  |  |
| **CNV** | **Cytoband** | **Type** | **Size (bp)** | **MAF** | **OR (95% CI)** | **β** | **SE** | ***P*** | **FDR *P*** |
|---|---|---|---|---|---|---|---|---|---|
| **EUR-like** |  |  |  |  |  |  |  |  |  |
| 2:238118075-238118809 | q37.3 | Del | 735 | 0.0029 | 3.38 (2.03-5.63) | 1.2177 | 0.2606 | 2.99E-06 | 0.0638 |
| 16:26721976-26722033 | p12.1 | Del | 58 | 0.0011 | 4.18 (2.28-7.65) | 1.4295 | 0.3089 | 3.70E-06 | 0.0638 |
| 2:91498696-91499277 | p11.2 | Dup | 582 | 0.0021 | 3.42 (2.05-5.71) | 1.2306 | 0.2608 | 2.38E-06 | 0.0638 |
| 22:47070265-47070423 | q13.31 | Del | 159 | 0.0008 | 7.41 (3.12-17.56) | 2.0026 | 0.4402 | 5.39E-06 | 0.06965 |
| **Trans-ancestry** |  |  |  |  |  |  |  |  |  |
| 16:26721976-26722033 | p12.1 | Del | 58 | 0.0011 | 2.93 (1.94-4.4153) | 1.0739 | 0.2098 | 3.06E-07 | 0.03049 |
| 2:88015229-88015399 | p11.2 | Del | 171 | 0.0003 | 14.99 (5.15-43.65) | 2.7073 | 0.5453 | 6.90E-07 | 0.03433 |
| **Meta-analysis of individual CNV** |  |  |  |  |  |  |  |  |  |
| **CNV** | **Cytoband** | **Type** | **Size (bp)** | **Weight** | **Z** | ***P*<sub>*meta*</sub>** | ***I*<sup>2</sup>** | **Het P** | **Gene Overlap** |
| 16:26721976-26722033 | p12.1 | Del | 58 | 12779.44 | 4.807 | 1.54E-06 | 33.9 | 0.2189 | ENSG00000260580 |

The 16p12.1 deletion is of particular interest because it was recovered independently in the trans-ancestry, EUR and meta-analytic results and falls within an established dosage-sensitive locus in which deletions are associated with developmental delay and with a broad range of psychiatric outcomes in both clinical and population-based samples (46–48). Its convergence across analytic levels, together with the absence of heterogeneity between ancestries, supports prioritising this locus for follow-up in larger AUD samples, although the event tested here is short and the association did not survive correction in any within-ancestry analysis.

Test statistics from the region-level analyses were well calibrated. For rCNV of all sizes, the λ was 1.03 in the trans-ancestry analysis and 1.05, 1.03 and 1.04 in the EUR, AFR and AMR samples, respectively. Restricting to rCNV of at least 10 kb yielded 1.05 in the trans-ancestry analysis, higher values in the EUR (1.21) and AFR (1.16) samples, and no inflation in the AMR sample (0.93); the smaller number of tests and the lower carrier frequency of large events make λ less stable in this category. The cross-ancestry meta-analyses gave λ = 1.05 for all CNV sizes and λ = 1.11 for rCNV of at least 10 kb, with λ_1000_ of 1.01 in both cases, indicating that the residual deviation from the null reflects sample size rather than uncontrolled population stratification. The current sample size may be underpowered to detect large rCNV associations, or alternatively, large rCNV may have little impact in AUD, while the functional role of small rCNV remains uncertain (49).

Taken together, all analyses converge on a consistent account. At the resolution of individual regions, no rCNV of at least 10 kb was associated with AUD after correction in any ancestry group, and the two deletions that reached FDR significance in the trans-ancestry analysis of all CNV sizes were both short events, at 16p12.1 and 2p11.2. At the level of burden, the associations that survived correction were almost exclusively inverse duplication effects recurring across biologically unrelated gene sets. The single deletion result that passed correction, together with the directionally consistent but uncorrected deletion signal in the xenobiotic response set, represents a possible candidate for an AUD-relevant effect. Both require replication in larger and more ancestrally diverse samples before firm conclusions can be drawn.

## Discussion

To our knowledge this is the first biobank-scale, WGS-based rCNV analysis of AUD using multi-level EHR phenotyping in 56,062 All of Us participants (5,410 cases, 50,652 controls). For rCNV ≥10 kb no genome-wide burden metric, or individual region was associated with AUD in any single or combined ancestry group after correction for multiple testing. Significant signals were modest and of two kinds. First, at the gene-set level, duplication burden and count were weakly associated with reduced risk of AUD across many gene sets, at both size thresholds, in the trans-ancestry analysis and the cross-ancestry meta-analysis but not within any single ancestry at ≥10 kb. Second, two short deletions (<200 bp) and genome-wide deletion count were associated with AUD only when rCNV of all sizes were included.

Duplications were associated with reduced AUD risk across biologically unrelated gene sets in the trans-ancestry analysis and the cross-ancestry meta-analysis, and three BrainSpan expression sets were significant under both the Stouffer and inverse-variance-weighted frameworks without heterogeneity. The cholesterol negative control was not significant for any metric (minimum FDR = 0.55), but its duplication count estimates fall in the same inverse direction as the significant sets, and it is underpowered to distinguish a global from a specific effect. The most simplest interpretation is that these results reflect a genome-wide property of duplication dosage, or residual confounding with duplication detection, rather than enrichment in AUD-relevant pathways.

The findings for deletions were narrower and more interpretable. Deletion burden was nominally elevated within the combined AUD/PAU gene set and the H-MAGMA brain chromatin-interaction set in the trans-ancestry analysis, deletion count within the Neurof_UnionStringent set survived correction, and deletion count within the abnormal behavioural response to xenobiotic set was nominally associated with AUD in the EUR, AMR and trans-ancestry analyses (Supplementary Tables 6 and 7). This is the pattern expected if deletions confer risk through haploinsufficiency of dosage-sensitive genes, as reported for other psychiatric phenotypes (21,27,50,51); however, with one exception, these results were not significant after correction.

The absence of FDR-significant regions is consistent with a highly polygenic disorder in which rare structural variation, if contributory, is distributed across many loci of small effect (11,24), and with limited power at the current case count. Two short deletions passed correction in the trans-ancestry analysis of rCNV of all sizes. The 58 bp deletion at 16p12.1 is the more compelling of the two: it also ranked first in the EUR-like and meta-analytic results, showed no between-ancestry heterogeneity, and falls within an established dosage-sensitive locus in which deletions are associated with developmental delay and a broad range of psychiatric outcomes (46–48). The EUR-like regions that approached significance also lie in or near genes of plausible neurobiological relevance, including *TBC1D22A* at 22q13.31 (52,53) and *ESPNL* at 2q37.3 (54,55). However all these individual rCNV are short, uncommon events, carrier counts are limiting, and replication in independent, adequately powered samples is required before they can be regarded as AUD loci.

While the current results suggest a modest contribution of rCNVs to AUD, they should be viewed in the context of several limitations. First, phenotyping of AUD from EHR in large-scale biobanks remains a developing area to address differences in healthcare utilization generally, and under-diagnosis and underreporting of AUD specifically. Second, CNV calls passing QC were available for only 56,062 participants in AUD case:control cohort (out of total CDRv8 srWGS 414,830 participants, of whom only 97,061 [23%] had srWGS-SV calls available). Additional verification of rCNVs across differing QC protocols is needed to confirm the robustness of small rCNVs previously undetectable in array data. Finally, the AFR and AMR samples remained underpowered and the EAS, CSA and MID groups were too small to analyse, so our results are still driven largely by EUR participants.

## Conclusion

rCNV detectable by short-read WGS do not appear to make a large contribution to AUD liability at this sample size. The 16p12.1 deletion and the xenobiotic-response deletion signal are the most promising candidates for follow-up, and distinguishing genuine pathway enrichment from global duplication load will require larger, more ancestrally diverse CNV datasets with harmonised calling and QC.

## Data Availability

All data produced in the present study are available upon reasonable request to the authors

https://github.com/saeedfv/AoU-CNV-Pipeline

## Data Availability

Individual-level phenotype and genetic data from the All of Us Research Program are available to approved researchers through the secure, Google Cloud-based All of Us Researcher Workbench. These data are not publicly available because of privacy and data-protection requirements. Eligible All of Us researchers can access individual-level phenotype, genetic, and inferred local ancestry data within the secure Researcher Workbench in accordance with the program’s data-access policies.

## Code Availability

All software tools and versions used in this study were as follows: PLINK 2.0, Python (v3.10.19), R (v4.4.2), bcftools (v1.24), bedtools (v2.29.1), and METAL (v2020-05-05). The complete code and analysis pipelines used in this study are publicly available in the GitHub repository: https://github.com/saeedfv/AoU-CNV-Pipeline.

## Acknowledgment

We gratefully acknowledge All of Us participants for their contributions, without whom this research would not have been possible. We also thank the National Institutes of Health’s All of Us Research Program for making available the participant data examined in this study.

## Supplementary Information

### Methods

#### Participants and phenotyping

A major challenge in studying AUD in biobanks is accurately phenotyping participants to select true cases and controls. AUD cases were defined based on the International Classification of Diseases, 9/10th Revision Clinical Modification (ICD-9/10-CM) diagnostic codes corresponding to SNOMED Clinical Terms (CT) concepts related to AUD and chronic alcohol-related conditions (Supplementary Table 1). We additionally identified 136 participants who lacked documented AUD-related diagnostic codes but had received prescriptions for acamprosate or disulfiram, both of which are specifically indicated for the treatment of AUD (1,2). Controls were selected from participants with no recorded AUD-related diagnoses or prescriptions and evidence of alcohol exposure based on the AoUs survey, thereby excluding individuals who reported lifetime abstinence.

Individuals who had been prescribed naltrexone in the absence of a corresponding AUD diagnosis (n = 2,695) were excluded to minimize potential misclassification arising from its use in the treatment of opioid use disorder (3). Following application of all phenotypic inclusion and exclusion criteria using EHR data in AoU, 29,485 individuals met the criteria for AUD and 483,907 individuals are classified as controls. Subsequent genomic quality-control procedures, including assessment of sex concordance and exclusion of ancestry outliers, yielded 22,385 AUD cases and 347,195 controls. Because CNV data were available for only a subset of participants with qualifying genomic data, the final CNV analysis included 56,062 participants (5410 cases and 50,652 controls) after applying CNV-specific quality-control criteria (Supplementary Figure 2).

#### Genetic Ancestry Assignment

We reassigned AoU participants to continental ancestry groups using the population grouping by Mahalanobis distance (POP-MaD) approach (4,5) with the same reference of merged 1000 Genomes Project and Human Genome Diversity Project (5,6) data as used by AoU. An individual was first assigned to the closest reference population defined by minimum Mahalanobis distance from the reference median calculated using the first 10 genetic principal components (PCs). Outliers in each initial group >3 S.D. from the median standardized within-group Mahalanobis distance were pruned. QC and LD-pruning was done within each assigned ancestry group. PCs were computed with FlashPCA2 (7) using the unrelated (<3^rd^ degree, estimated using KING (8)) subset of assigned ancestry groups. Within-ancestry related individuals and ancestry outliers were then projected onto the PCs. For trans-ancestry PCs, the union of unrelated individuals from each assigned ancestry group was used to compute PCs onto which remaining individuals were projected. We identified European-like (EUR), African-like (AFR), Admixed American-like (AMR), East Asian-like (EAS), Central and South Asian-like (CSA), Middle Eastern-like (MID), and Oceania-like (OCE) ancestries. All models included the first 10 within-ancestry PCs, age, and sex at birth. Because only a subset of AoU participants have CNV calls, the subsamples of EAS, CSA, MID, and OCE ancestries were all less than 1000 individuals, so we included only EUR, AFR and AMR groups (Table 1) for within-ancestry analyses (Supplementary Figure 3).

#### All of Us WGS and CNV detection

This study used data from the All of Us (AoU) Curated Data Repository (CDR) release C2024Q3R3 (“CDRv8”), which includes short-read whole-genome sequencing (srWGS) data for 414,830 participants, produced at the Broad Institute, Baylor College of Medicine, and the University of Washington using Illumina NovaSeq 6000 sequencers. Data processing pipelines and data QC procedures (9) (https://support.researchallofus.org/hc/en-us/articles/29390274413716-All-of-Us-Genomic-Quality-Report) have been published. Our analyses were based on the srWGS structural variant (SV) callset, which comprises 97,061 participants and is an updated version of the CDRv7 off-cycle srWGS SV release (97,940 samples). For CDRv8, variant calling was not rerun; instead, dropped samples were removed and additional refinements were applied. Notably, the SV callset was generated from CRAMs aligned with DRAGEN v3.4.12, whereas other srWGS data in CDRv8 used DRAGEN v3.7.8. All samples followed the Consistency across Genome Centers and Single Sample QC processes in the srWGS QC pipeline prior to SV calling. SVs were called with GATK-SV using the previously described pipeline (10), which combines Manta and Wham (paired-end/split-read evidence), MELT (mobile elements), and GATK-gCNV and cn.MOPS (read-depth CNVs). During callset generation, samples were removed for insert size outside 320–700 bp, assessed using Picard (CollectInsertSizeMetrics within GATK’s CollectMultipleMetrics; N = 30); contamination ≥1% (N = 296); whole-genome dosage (WGD) scores >6× MAD, because coverage variability affects CNV calling (N = 1,337); >500 non-diploid 1 Mb bins (N = 1,508); and outlier SV counts across ≥30 categories or abnormal chromosome ploidy (copy ratio outside 1.8–2.3; N = 61). We then applied additional filters. Samples flagged for srWGS QC issues (N = 4,044) were excluded prior to all further filtering. We compared the DRAGEN sex-chromosome constitution against self-reported sex assigned at birth, retaining only individuals who self-reported “male” or “female” and had a matching constitution (XY or XX), and removed samples with potential sex-chromosome aneuploidy. Related individuals were excluded using the AoU kinship matrix, so that only unrelated individuals were retained. Finally, we restricted analyses to rare (MAF < 1%) deletions and duplications and excluded all other SV types.

#### Individual rCNV filtering

CNV region-based association testing (duplications and deletions analyzed separately) was performed in PLINK 2.0 using Firth’s logistic regression, adjusting for age, sex, and PCs 1-10. Because regions observed in only a few individuals yield unstable association metrics and inflated test statistics, inference was restricted to regions with a minor allele count (MAC) > 20; regions below this threshold were excluded prior to multiple-testing correction. Region-level p-values for the retained tests were corrected using the Benjamini–Hochberg false discovery rate, and associations passing FDR < [0.05] were reported. The genomic inflation factor was evaluated on the MAC-filtered test set. A total of 863,195 CNVs (DELs plus DUPs) are excluded due to MAC <= 20. To comply with the All of Us Research Program Data and Statistics Dissemination Policy, which prohibits reporting statistics derived from ≤ 20 individuals, we further confirmed that every reported region was supported by more than 20 unique carrier individuals, counting each heterozygous or homozygous carrier once [https://support.researchallofus.org/hc/en-us/articles/360043016291-How-to-comply-with-the-All-of-Us-Data-and-Statistics-Dissemination-Policy].

#### CNV burden analysis

CNV burden was assessed at both the genome-wide and gene-set levels, following a previously published analytic framework (11). CNV burden was quantified using multiple complementary metrics, including: (1) the cumulative length in megabases (Mb) of all rare duplications and deletions analyzed separately; (2) the number of rare duplications and deletions considered separately; and (3) the aggregate length of all duplications and deletions combined, and evaluated for association with AUD using Firth’s logistic regression. We initially analyzed CNV >10 Kb in size due to their greater potential for pathogenicity, and then analyzed rCNV of any size. We conducted both genome-wide and gene-set burden analyses within ancestry for the three largest ancestry groups (EUR, AFR, AMR), we performed meta-analysis of the within-ancestry results, and we performed a trans-ancestry analysis of all samples pooled together. Within-ancestry meta-analysis and trans-ancestry analysis have different strengths and limitations. Because CNV boundaries vary by ancestry, meta-analysis of within-ancestry summary statistics across groups controls for possible population stratification by keeping null models calibrated within each ancestral group. However, AoU is a population cohort and, like the US population, displays substantial admixture. Trans-ancestry analysis increases both sample size and power, and appears to better address this admixture.

#### Genome-wide burden analysis

We conducted whole-genome rCNV burden analyses for both deletions and duplications across count, length, and the cumulative length metrics using Firth’s logistic regression, adjusted for the first ten genetic principal components to control for population stratification. We used three covariate models for genome-wide burden tests: 1) ten PCs only and 2) ten PCs, age, sex and 3) ten PCs, age, sex, total rCNV count, and average rCNV length. (Supplementary Figure 4). Effect estimates were broadly consistent across models. Following previous studies (35,42) we report Model 3 as the primary model in the main text because it is the most conservative, additionally accounting for individual differences in global rCNV load.

#### Gene-set burden analysis

At the gene-set level, analyses quantified the extent to which rCNV overlapped individual genes and predefined gene sets. To map genes to each identified CNV to calculate burden in Mb, we used the GENCODE Release 46 (GRCh38.p14) (12) genic intervals. Each CNV was assigned to any genes sharing at least one base pair of overlap (11).

We also conducted gene-set rCNV analyses of both deletions and duplications across count (representing the number of genes within the set overlapped by CNV in each individual), length and the cumulative length metrics using Firth’s logistic regression. Gene-set burden analysis included 5 AUD/problematic alcohol use (PAU) related gene sets from a previous PAU study (13) plus a combination of all five sets, 146 abnormal behavior gene sets derived from mouse mutants with central nervous system (CNS) phenotypes (14–16), 73 curated sets studied for CNV from PGC PTSD study and one negative control cholesterol gene-set, previously shown to have no genetic association with PAU (13). For each gene, the total number of overlapping CNV and the extent of base pair overlap were calculated. Gene burden was encoded as a binary variable, where individuals were assigned a value of 1 if at least one CNV overlapped the gene and 0 otherwise.

We adjusted for average rCNV length and total rCNV count to reduce confounding by background genomic instability. Total rCNV count accounts for differences in the rate of rCNV formation between cases and controls, while average rCNV length accounts for differences in the size of the events themselves (17).

Together, these covariates capture genome-wide rCNV burden, so that associations reflect the specific loci of interest rather than a global tendency toward structural variation. We evaluated multiple covariate models to identify the optimal predictors of CNV burden [Supplementary method]; however, our primary model followed previously established frameworks (11) and tested the five gene-set level burden metrics: deletion/duplication count, deletion/duplication burden length, and total burden length within each gene set. For each metric, we fitted Firth’s logistic regression model of the following form, adjusting for genome-wide average rCNV length, genome-wide total rCNV length, the ten within-ancestry genetic PCs, age, and sex.

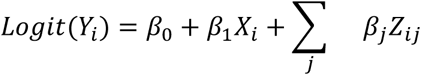

#### Cross-ancestry burden meta-analysis

Ancestry-specific *β* estimates and their SEs were combined across ancestries (EUR, AFR and AMR) using fixed-effects meta-analysis under two complementary frameworks. First, a sample-size–weighted Stouffer’s Z (18) was computed because it weights each ancestry by the square root of its sample size (*w_i_* = √*N_i_*), making it more robust to imbalance in cohort size and to instability in per-ancestry standard-error estimates. Stouffer’ *Z_meta_* computed as

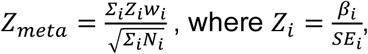

where *i* indexes ancestry and *N_i_* is the corresponding cohort sample size, and a two-sided meta-analysis *P* value (*P*_Meta_) was obtained from the standard normal distribution as *P_meta_* = 2 × [1 − *Φ*(|*Z_meta_*|)]. Second, an inverse-variance–weighted (IVW) estimate was obtained as

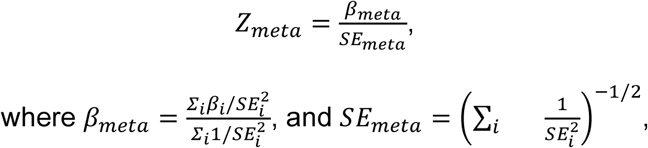

with a corresponding pooled odds ratio (OR) and 95% confidence interval derived by exponentiating *β_meta_* and its interval. Two-sided *P* values were computed from the standard normal distribution for both frameworks.

#### Individual rCNV analysis

rCNV associations with AUD were evaluated using Firth’s logistic regression to account for sparse data and low-frequency events. Subsequent ancestry-stratified analyses were conducted within the EUR, AFR, and AMR groups. The CSA, EAS, and MID groups were excluded from ancestry-specific analyses because limited sample sizes reduced statistical power for rare CNV association testing. We also performed an exploratory analysis in a trans-ancestry framework using all QC-passed samples from all ancestry groups (including EUR, AMR, AFR, and adding CSA, EAS, and MID) and using transancestry PCs for assignment, as described above. Within-and trans-ancestry ancestry association analyses were performed using PLINK 2.0 (Chang et al., 2015). For each model, age at blood collection, sex, and the first 10 within-ancestry PCs (for within-ancestry analyses) or trans-ancestry PCs (for trans-ancestry analyses) were utilized as covariates.

Associations between rCNV and AUD were evaluated using p-values (*P*), regression coefficients (*β*), OR, and standard errors (SE) derived from the regression models. We analyzed CNV regions with two filters of ≥10 kb and all CNV sizes

#### Individual rCNV meta-analysis

Association results for individual CNV regions in the EUR, AFR, and AMR ancestry groups were meta-analyzed using METAL (19) with the sample-size weighted scheme (SCHEME SAMPLESIZE), which combines per-ancestry *P* values into a weighted Stouffer’s *Z*, weighting each study by the square root of its sample size and incorporating the direction of effect. This matches the sample-size weighted framework used for the gene-set burden meta-analysis above. To account for imbalanced case-control ratios, the effective sample size (*N_eff_*) was used as the sample size for each study.

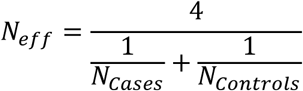

Genomic control correction was applied. Between-ancestry heterogeneity was assessed using Cochran’s *Q* and *I*² (ANALYZE HETEROGENEITY). Significance was assessed after FDR correction across the *n* regions tested with CNV size thresholds (all CNV sizes and ≥10kb CNV).

### Multiple test correction

Statistical significance was determined using the Benjamini–Hochberg false discovery rate (FDR), with an adjusted p-value threshold of <0.05 applied within each family of tests and adjusting within ancestry group and CNV size filters.

### Genomic inflation evaluation

To assess potential inflation of association statistics, we first calculated the genomic inflation factor (λ), defined as the ratio of the median observed chi-square association test statistic to the median expected under the null hypothesis of no association. Because λ is influenced by sample size, we additionally calculated the standardized genomic inflation factor λ₁₀₀₀ (20), which adjusts λ to an equivalent study with 1,000 cases and 1,000 controls using the following formula.

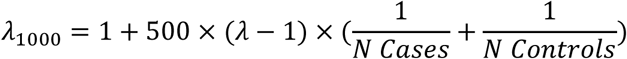

Values greater than 1 for either metric indicate inflation of the test statistics, whereas values close to 1 suggest adequate control of population stratification and other confounding factors (21).

### Results

#### Gene sets Burden

The following table shows the description of gene sets:

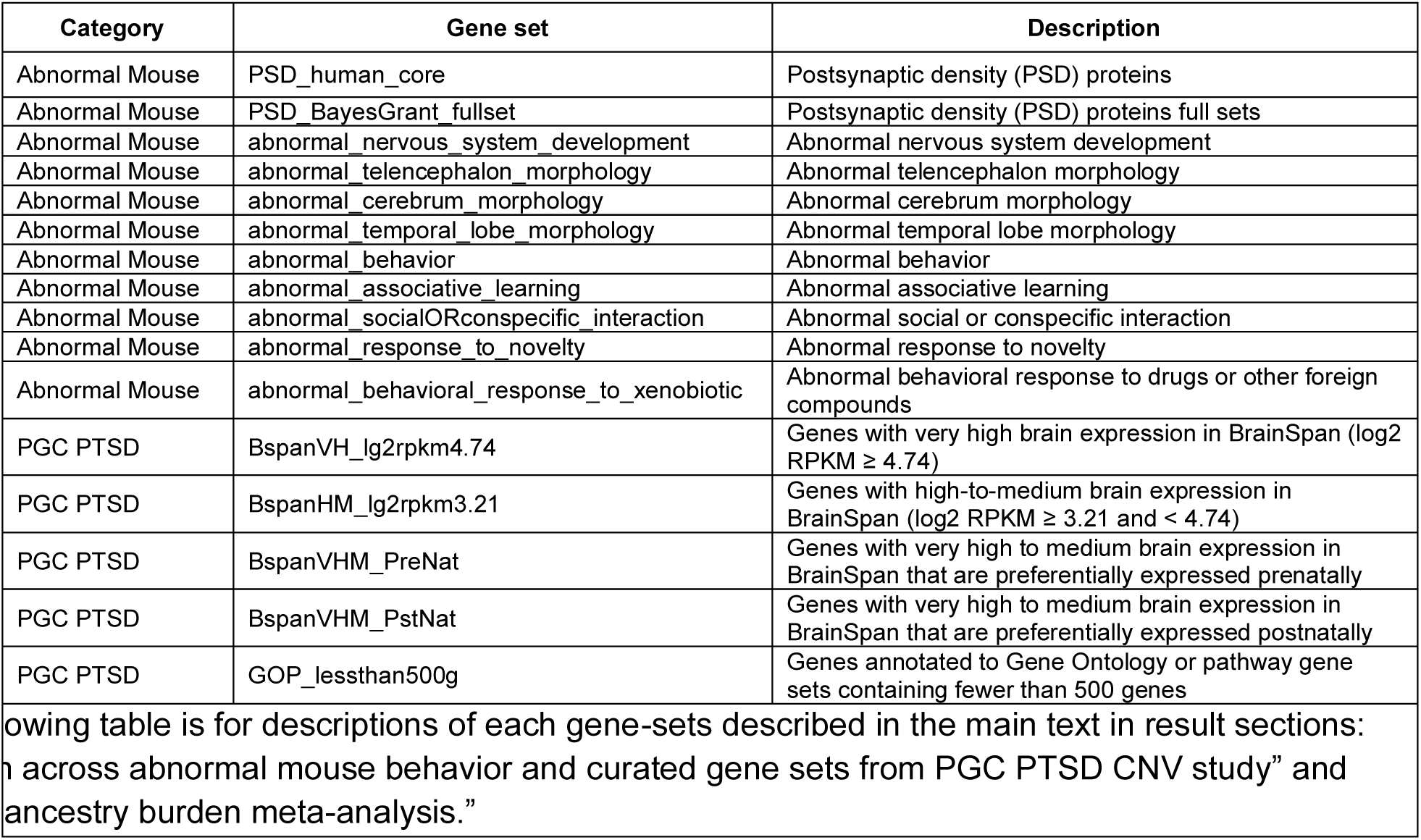

| Category | Gene set | Description |
|---|---|---|
| Abnormal Mouse | PSD_human_core | Postsynaptic density (PSD) proteins |
| Abnormal Mouse | PSD_BayesGrant_fullset | Postsynaptic density (PSD) proteins full sets |
| Abnormal Mouse | abnormal_nervous_system_development | Abnormal nervous system development |
| Abnormal Mouse | abnormal_telencephalon_morphology | Abnormal telencephalon morphology |
| Abnormal Mouse | abnormal_cerebrum_morphology | Abnormal cerebrum morphology |
| Abnormal Mouse | abnormal_temporal_lobe_morphology | Abnormal temporal lobe morphology |
| Abnormal Mouse | abnormal_behavior | Abnormal behavior |
| Abnormal Mouse | abnormal_associative_learning | Abnormal associative learning |
| Abnormal Mouse | abnormal_socialORconspecific_interaction | Abnormal social or conspecific interaction |
| Abnormal Mouse | abnormal_response_to_novelty | Abnormal response to novelty |
| Abnormal Mouse | abnormal_behavioral_response_to_xenobiotic | Abnormal behavioral response to drugs or other foreign compounds |
| PGC PTSD | BspanVH_lg2rpkm4.74 | Genes with very high brain expression in BrainSpan (log2 RPKM ≥ 4.74) |
| PGC PTSD | BspanHM_lg2rpkm3.21 | Genes with high-to-medium brain expression in BrainSpan (log2 RPKM ≥ 3.21 and < 4.74) |
| PGC PTSD | BspanVHM_PreNat | Genes with very high to medium brain expression in BrainSpan that are preferentially expressed prenatally |
| PGC PTSD | BspanVHM_PstNat | Genes with very high to medium brain expression in BrainSpan that are preferentially expressed postnatally |
| PGC PTSD | GOP_lessthan500g | Genes annotated to Gene Ontology or pathway gene sets containing fewer than 500 genes |

[illegible]owing table is for descriptions of each gene-sets described in the main text in result sections: [illegible] across abnormal mouse behavior and curated gene sets from PGC PTSD CNV study” and [illegible]ancestry burden meta-analysis.”

#### Association of individual rCNV with AUD

No rCNV region of at least 10 kb was associated with AUD after FDR correction in the EUR, AFR or AMR samples, in the trans-ancestry analysis, or in the cross-ancestry meta-analysis. When rCNV of any size were considered, two deletions reached FDR significance in the trans-ancestry analysis. A 58 bp deletion at 16p12.1 overlapping *ENSG00000260580* gave the smallest *P* value genome-wide (OR = 2.93, 95% CI 1.94 to 4.42, P = 3.06 × 10-7, FDR = 0.030), and a 171 bp intergenic deletion at 2p11.2 was also significant (OR = 14.99, 95% CI 5.15 to 43.65, *P* = 6.90 × 10-7, FDR = 0.034). The 16p12.1 deletion was likewise the top-ranked region in the EUR sample (OR = 4.18, 95% CI 2.28 to 7.65, *P* = 3.70 × 10-6, FDR = 0.064), where three further regions approached significance: a 582 bp intergenic duplication at 2p11.2 (OR = 3.42, 95% CI 2.05 to 5.71, *P* = 2.38 × 10-6, FDR = 0.064), a 735 bp deletion at 2q37.3 overlapping *ESPNL* (OR = 3.38, 95% CI 2.03 to 5.63, *P* = 2.99 × 10-6, FDR = 0.064) and a 159 bp deletion at 22q13.31 overlapping *TBC1D22A* (OR = 7.41, 95% CI 3.13 to 17.56, *P* = 5.39 × 10-6, FDR = 0.070). No region survived FDR correction in the AFR-like or AMR-like samples, possibly consistent with their smaller case counts. In the cross-ancestry meta-analysis of the three groups, the 16p12.1 deletion again ranked first (Z = 4.81, *P* = 1.54 × 10⁻⁶, FDR = 0.10), although no region passed correction (FDR < 0.05). There was no significant between-ancestry heterogeneity at this locus (I² = 33.9%, P heterogeneity = 0.22). All implicated events were very short and uncommon, and each conferred a roughly three-fold to fifteen-fold increase in the odds of AUD among carriers.

#### Individual CNV region and meta analysis results

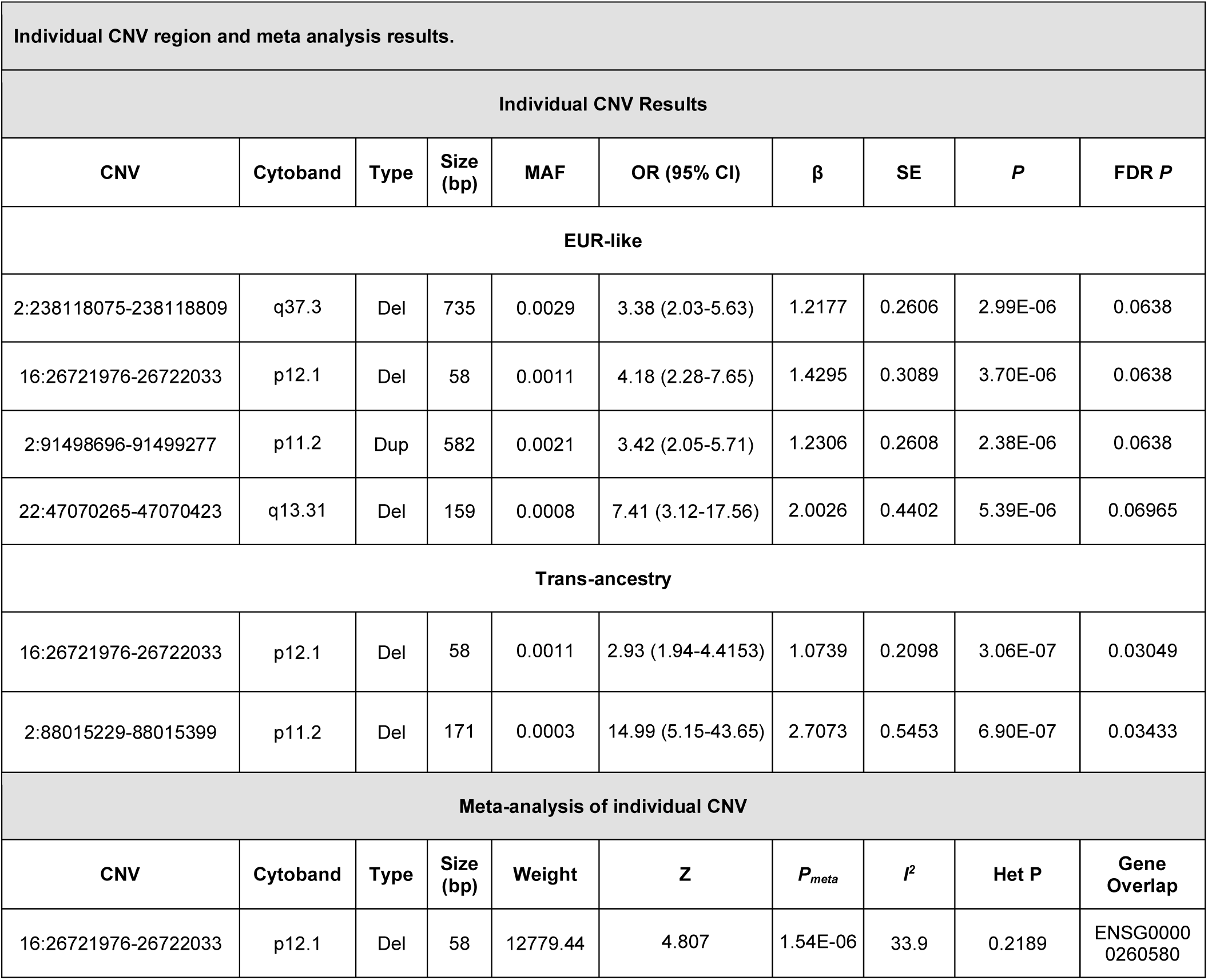

| **Individual CNV Results** |  |  |  |  |  |  |  |  |  |
| **CNV** | **Cytoband** | **Type** | **Size (bp)** | **MAF** | **OR (95% CI)** | **β** | **SE** | ***P*** | **FDR *P*** |
|---|---|---|---|---|---|---|---|---|---|
| **EUR-like** |  |  |  |  |  |  |  |  |  |
| 2:238118075-238118809 | q37.3 | Del | 735 | 0.0029 | 3.38 (2.03-5.63) | 1.2177 | 0.2606 | 2.99E-06 | 0.0638 |
| 16:26721976-26722033 | p12.1 | Del | 58 | 0.0011 | 4.18 (2.28-7.65) | 1.4295 | 0.3089 | 3.70E-06 | 0.0638 |
| 2:91498696-91499277 | p11.2 | Dup | 582 | 0.0021 | 3.42 (2.05-5.71) | 1.2306 | 0.2608 | 2.38E-06 | 0.0638 |
| 22:47070265-47070423 | q13.31 | Del | 159 | 0.0008 | 7.41 (3.12-17.56) | 2.0026 | 0.4402 | 5.39E-06 | 0.06965 |
| **Trans-ancestry** |  |  |  |  |  |  |  |  |  |
| 16:26721976-26722033 | p12.1 | Del | 58 | 0.0011 | 2.93 (1.94-4.4153) | 1.0739 | 0.2098 | 3.06E-07 | 0.03049 |
| 2:88015229-88015399 | p11.2 | Del | 171 | 0.0003 | 14.99 (5.15-43.65) | 2.7073 | 0.5453 | 6.90E-07 | 0.03433 |
| **Meta-analysis of individual CNV** |  |  |  |  |  |  |  |  |  |
| **CNV** | **Cytoband** | **Type** | **Size (bp)** | **Weight** | **Z** | ***P*<sub>*meta*</sub>** | ***I*<sup>2</sup>** | **Het P** | **Gene Overlap** |
| 16:26721976-26722033 | p12.1 | Del | 58 | 12779.44 | 4.807 | 1.54E-06 | 33.9 | 0.2189 | ENSG00000260580 |

The 16p12.1 deletion is of particular interest because it was recovered independently in the trans-ancestry, EUR and meta-analytic results and falls within an established dosage-sensitive locus in which deletions are associated with developmental delay and with a broad range of psychiatric outcomes in both clinical and population-based samples (22–24). Its convergence across analytic levels, together with the absence of heterogeneity between ancestries, supports prioritising this locus for follow-up in larger AUD samples, although the event tested here is short and the association did not survive correction in any within-ancestry analysis.

Test statistics from the region-level analyses were well calibrated. For rCNV of all sizes, the λ was 1.03 in the trans-ancestry analysis and 1.05, 1.03 and 1.04 in the EUR, AFR and AMR samples, respectively. Restricting to rCNV of at least 10 kb yielded 1.05 in the trans-ancestry analysis, higher values in the EUR (1.21) and AFR (1.16) samples, and no inflation in the AMR sample (0.93); the smaller number of tests and the lower carrier frequency of large events make λ less stable in this category. The cross-ancestry meta-analyses gave λ = 1.05 for all CNV sizes and λ = 1.11 for rCNV of at least 10 kb, with λ_1000_ of 1.01 in both cases, indicating that the residual deviation from the null reflects sample size rather than uncontrolled population stratification (see the table below for the complete λ values).

#### Genomic inflation factors for individual rCNV association analyses

| **Analysis** | **All sizes: λ** | **All sizes: λ<sub>1000</sub>** | **≥10 kb: λ** | **≥10 kb: λ<sub>1000</sub>** |
|---|---|---|---|---|
| EUR | 1.05 | 1.01 | 1.21 | 1.05 |
| AFR | 1.03 | 1.01 | 1.16 | 1.08 |
| AMR | 1.04 | 1.03 | 0.93 | 0.95 |
| Trans-ancestry | 1.03 | 1.00 | 1.05 | 1.01 |
| Cross-ancestry meta-analysis | 1.05 | 1.01 | 1.11 | 1.01 |

The current sample size may be underpowered to detect large rCNV associations, or alternatively, large rCNV may have little impact in AUD, while the functional role of small rCNV remains uncertain (25).

### Supplementary Figures

**Supplementary figure 1.**
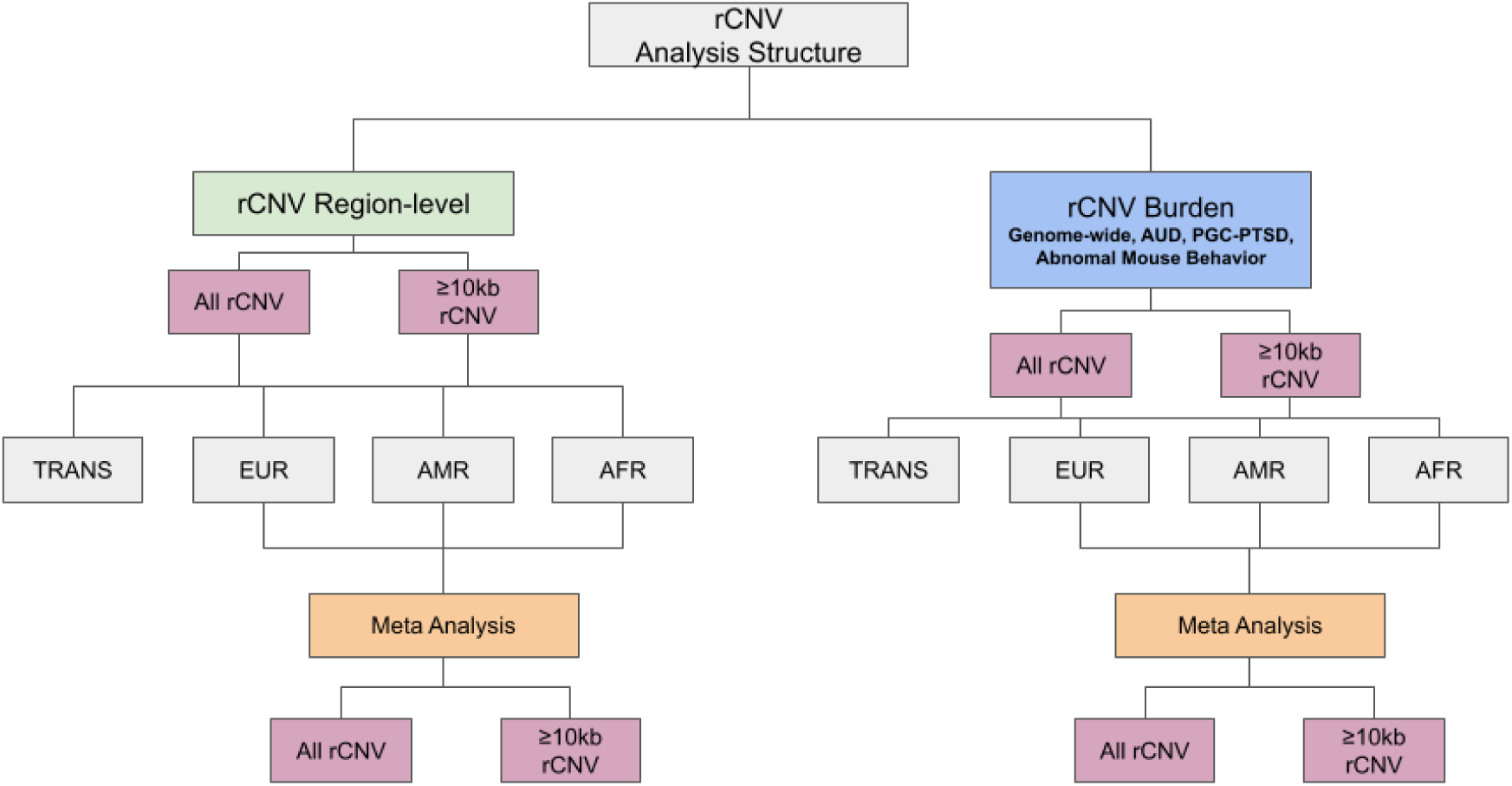
Overview of the rare copy number variant (rCNV) analysis structure. Two complementary analyses were performed: region-level rCNV association and rCNV burden (genome-wide and within AUD, PGC-PTSD, and abnormal mouse behavior gene sets). Each was run for all rCNVs and for rCNVs ≥10 kb, in the trans-ancestry (TRANS, pooled samples) sample and separately in European (EUR), admixed American (AMR), and African (AFR) ancestry groups. Ancestry-specific results were then combined by meta-analysis for both rCNV size categories.

**Supplementary figure 2.**
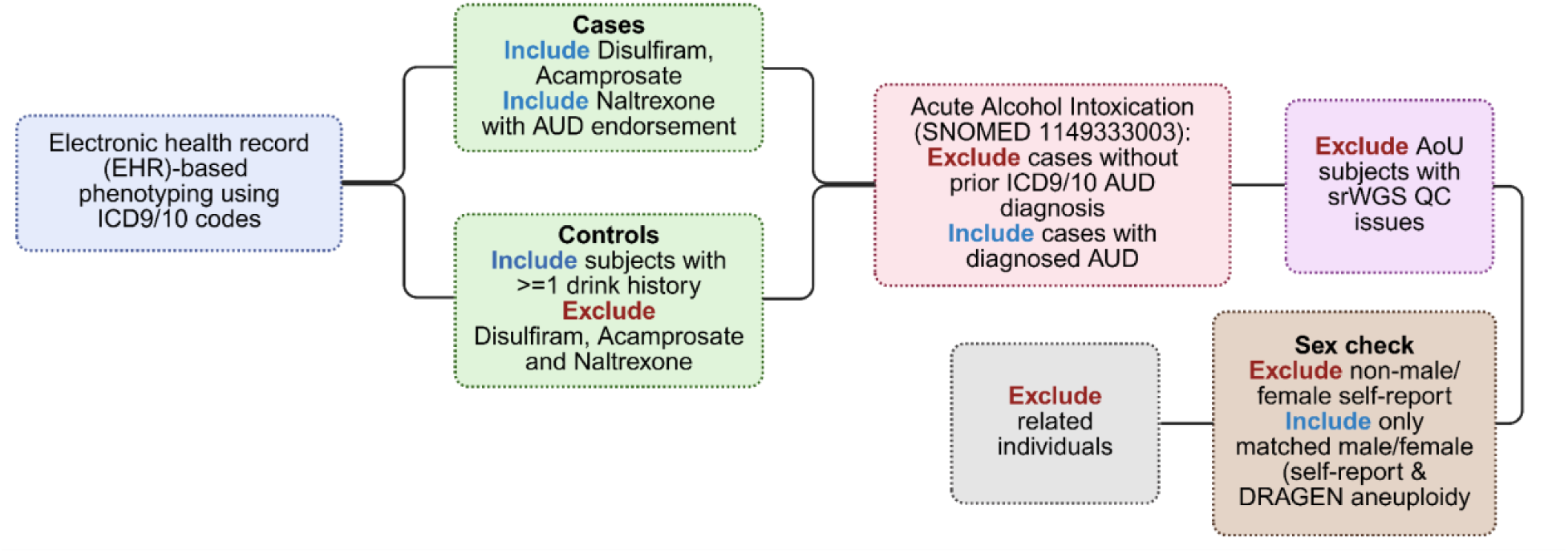
Cohort selection and quality-control workflow for alcohol use disorder (AUD) cases and controls in the All of Us (AoU) Research Program. Participants were phenotyped from electronic health records (EHR) using ICD-9/10 codes. Cases additionally included individuals prescribed disulfiram or acamprosate, or naltrexone with an accompanying AUD endorsement. Controls were restricted to individuals reporting at least one lifetime drink and no exposure to disulfiram, acamprosate, or naltrexone. Individuals with acute alcohol intoxication (SNOMED 1149333003) were retained as cases only if they had a prior ICD-9/10 AUD diagnosis. Participants were then excluded sequentially for short-read whole-genome sequencing (srWGS) quality-control failures, non-male/female self-reported sex or discordance between self-reported sex and DRAGEN-inferred sex chromosome ploidy, and relatedness.

**Supplementary figure 3.**
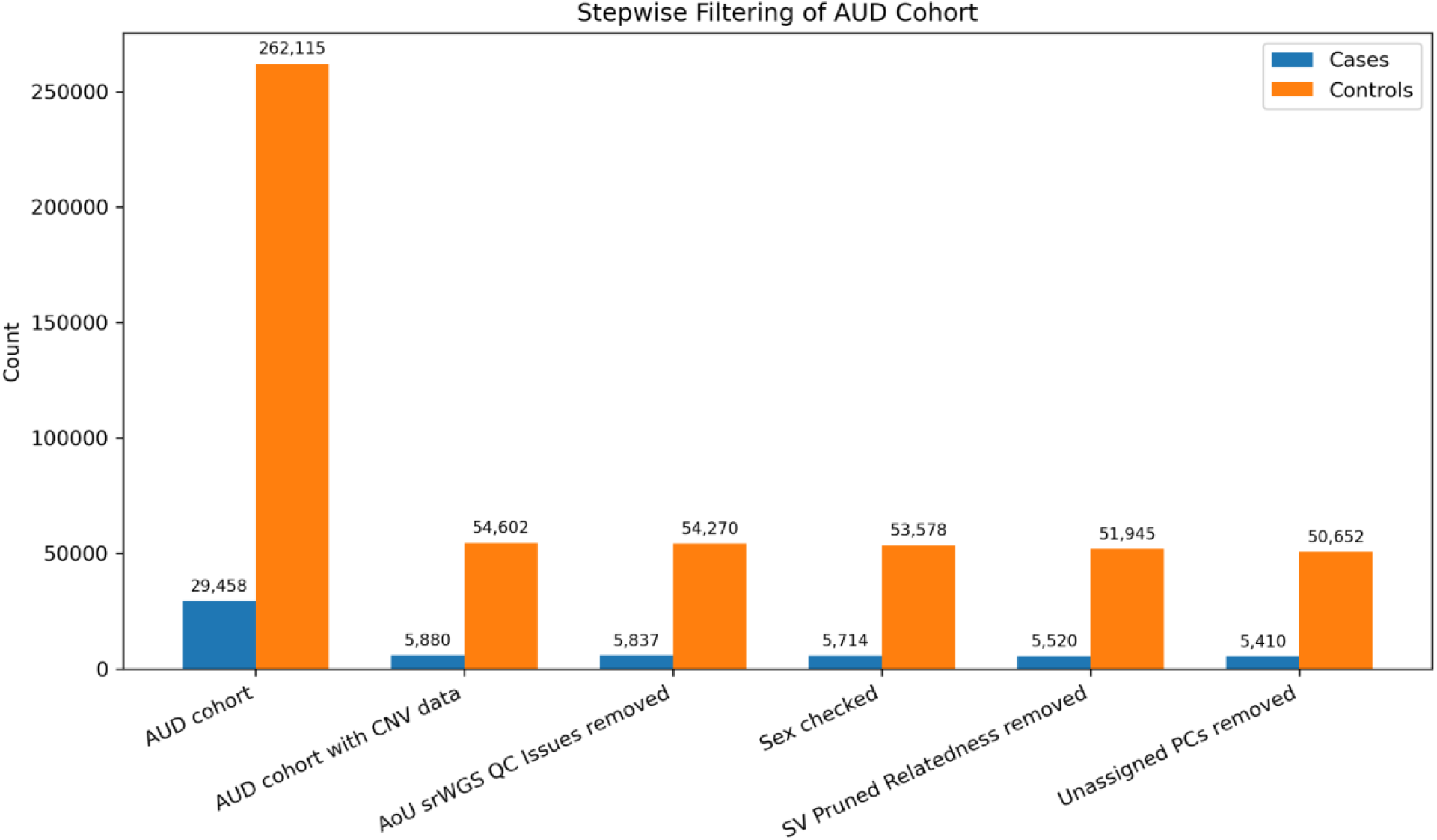
Stepwise sample filtering of the AUD cohort in All of Us. Number of cases (blue) and controls (orange) retained after each filtering step: (1) the initial EHR-defined AUD cohort (29,458 cases; 262,115 controls), (2) restriction to participants with CNV data, (3) removal of flagged srWGS quality-control issues, (4) sex concordance check, (5) removal of related individuals, and (6) removal of participants with unassigned genetic ancestry based on principal components (PCs). The final analytic cohort included 5,410 cases and 50,652 controls.

**Supplementary Figure 4.**
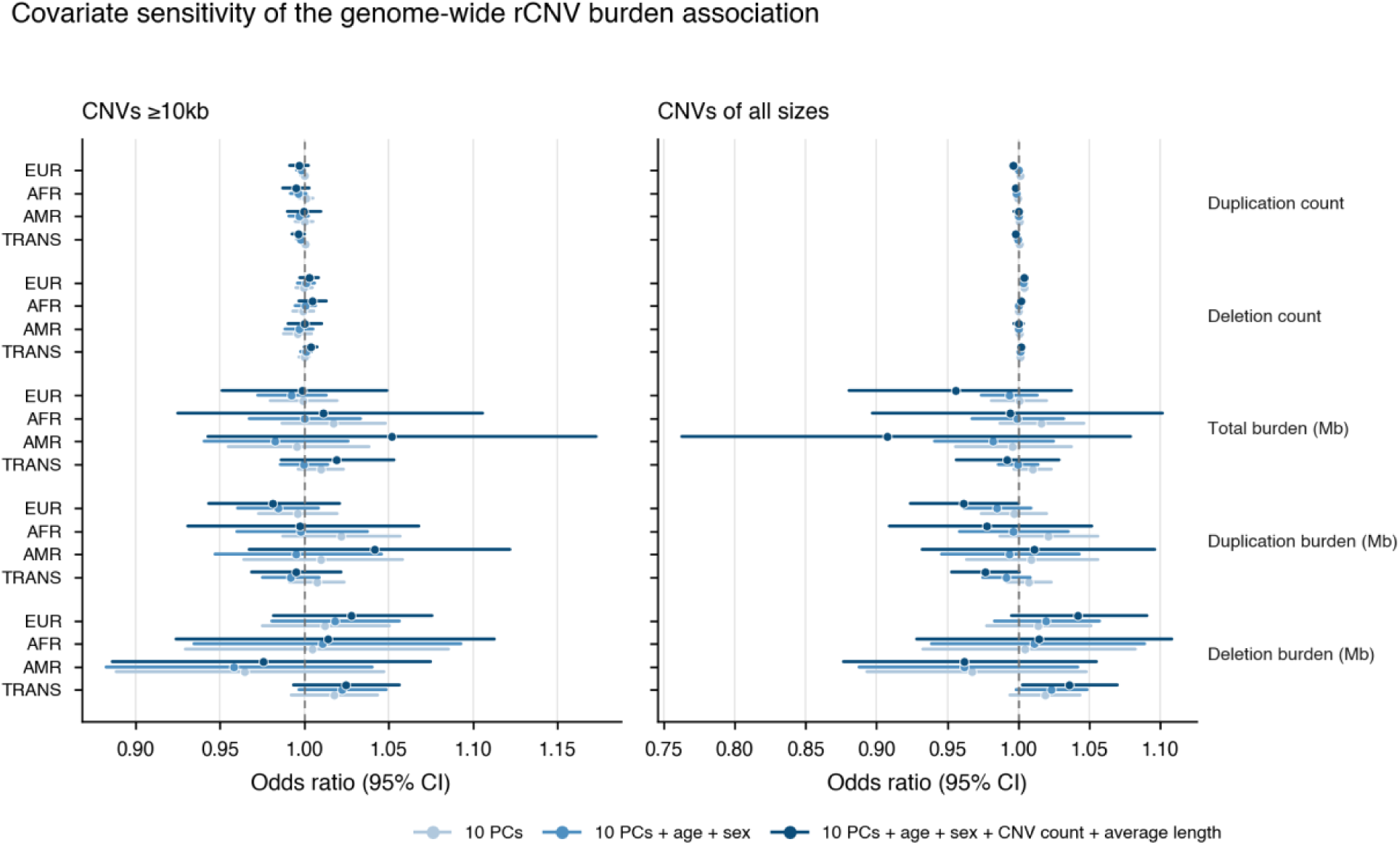
Covariate sensitivity of the genome-wide rCNV burden association with AUD. Odds ratios and 95% confidence intervals for five rCNV burden metrics (duplication count, deletion count, and total, duplication, and deletion burden in Mb) are shown for rCNVs ≥10 kb (left) and rCNVs of all sizes (right), in European (EUR), African (AFR), and admixed American (AMR) ancestry groups and the trans-ancestry (TRANS) sample. Three nested covariate models are compared: 10 principal components (PCs) only; 10 PCs plus age and sex; and 10 PCs, age, sex, CNV count, and average CNV length. The dashed line marks an odds ratio of 1. Estimates were broadly consistent across models, with wider confidence intervals for the Mb-based burden metrics in the fully adjusted model.

**Supplementary Figure 5.**
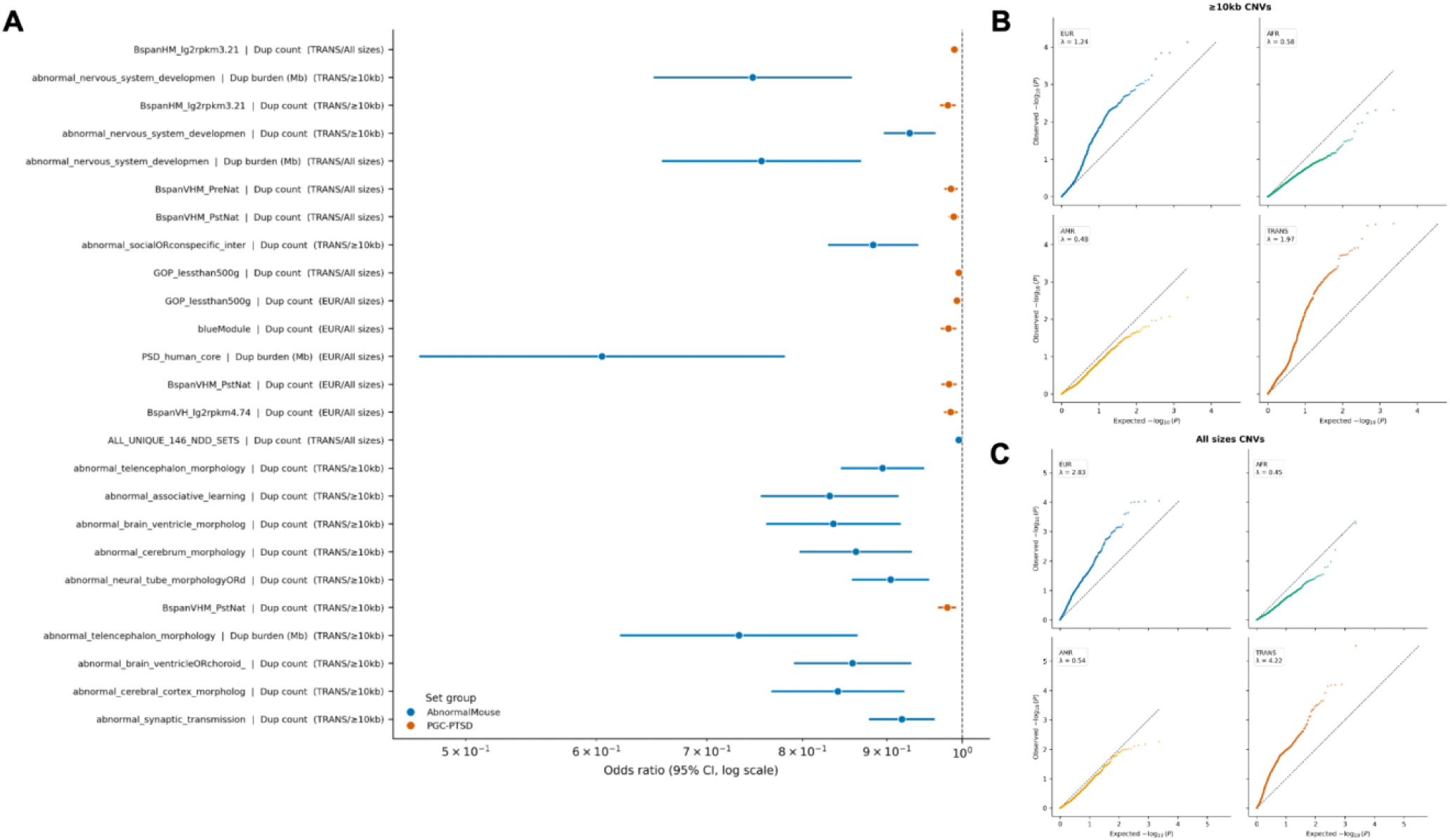
Gene-set rCNV burden associations with AUD. **(A)** Odds ratios and 95% confidence intervals (log scale) for gene-set burden associations (FDR < 0.05). Each row shows the gene set, burden metric (duplication count or duplication burden in Mb), and analysis category (ancestry group/rCNV size category). Colors indicate gene-set group (blue, abnormal mouse phenotype; orange, PGC-PTSD). The dashed line marks an odds ratio of 1. **(B, C)** Quantile–quantile plots of observed versus expected −log10(*P*) values from gene-set burden tests for rCNVs ≥10 kb (B) and rCNVs of all sizes (C), shown separately for European-like (EUR), African-like (AFR), and admixed American-like (AMR) ancestry groups and the trans-ancestry (TRANS) sample. λ is the genomic inflation factor.

**Supplementary Figure 6.**
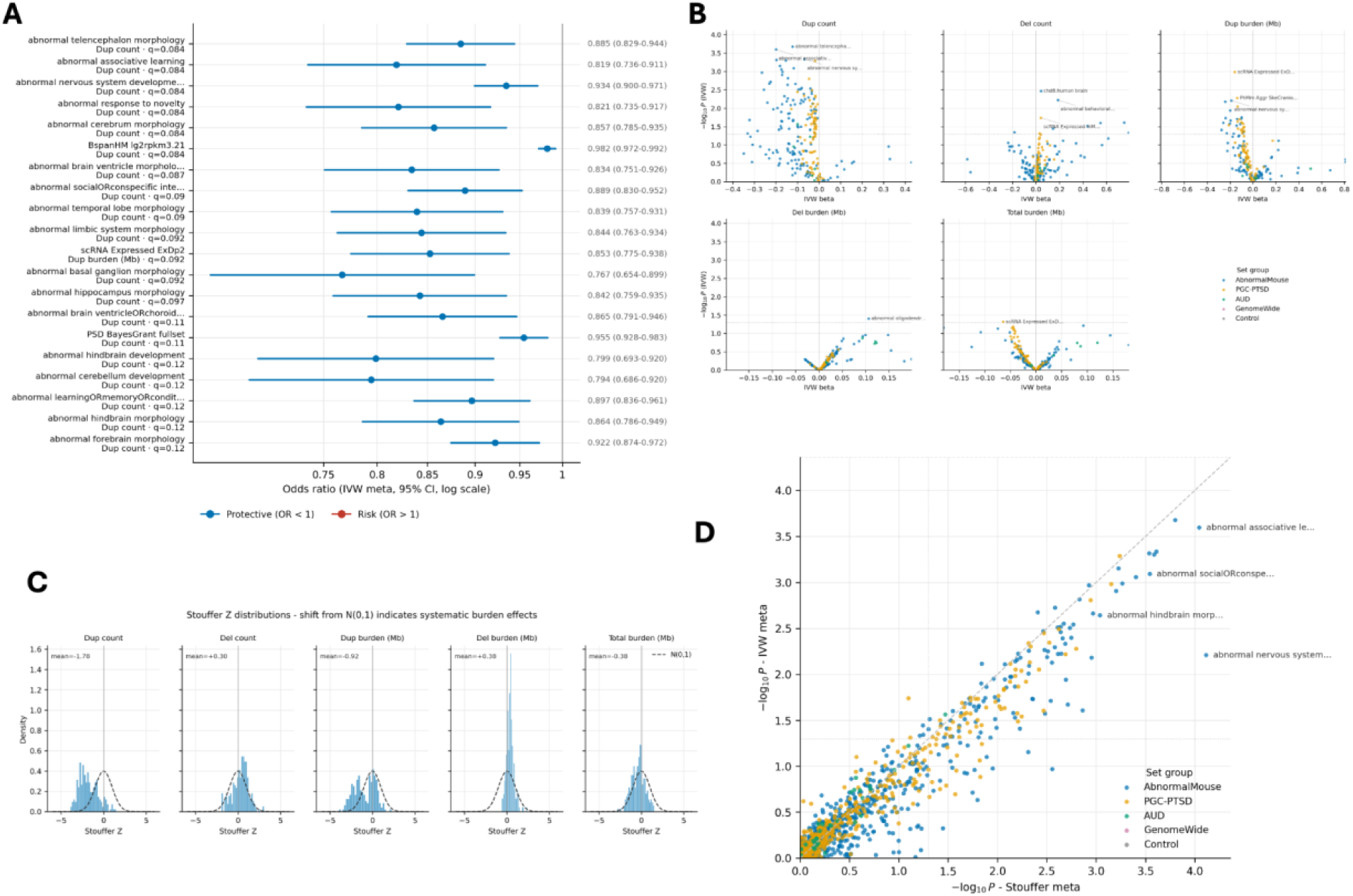
Cross-ancestry meta-analysis of gene-set rCNV burden in AUD (rCNVs ≥10 kb). **(A)** Odds ratios and 95% confidence intervals (log scale) for the 20 gene set–burden metric pairs with the smallest inverse-variance-weighted (IVW) meta-analysis *P*-values across European, African, and admixed American ancestry groups. Row labels give the gene set, burden metric, and FDR *q*-value; estimates are listed at right. Blue indicates odds ratios below 1 and red indicates odds ratios above 1. **(B)** Volcano plots of IVW effect size (beta) against −log10(*P*) for each burden metric: duplication count, deletion count, and duplication, deletion, and total burden (Mb). Points are colored by gene-set group. The dotted line marks *P* = 0.05; the most significant sets are labeled. **(C)** Distribution of Stouffer meta-analysis Z-scores across gene sets for each burden metric, compared with the standard normal distribution (dashed curve); the mean Z-score is shown in each panel. **(D)** Comparison of −log10(*P*) from Stouffer and IVW meta-analyses across all gene set–burden metric tests, colored by gene-set group. The diagonal indicates equal *P*-values; the dotted lines mark *P* = 0.05 and the dashed line marks the Bonferroni threshold. All models were adjusted for ten principal components, age, sex, total rCNV count, and average rCNV length. No association reached FDR *q* < 0.05 in the IVW meta-analysis.

**Supplementary Figure 7.**
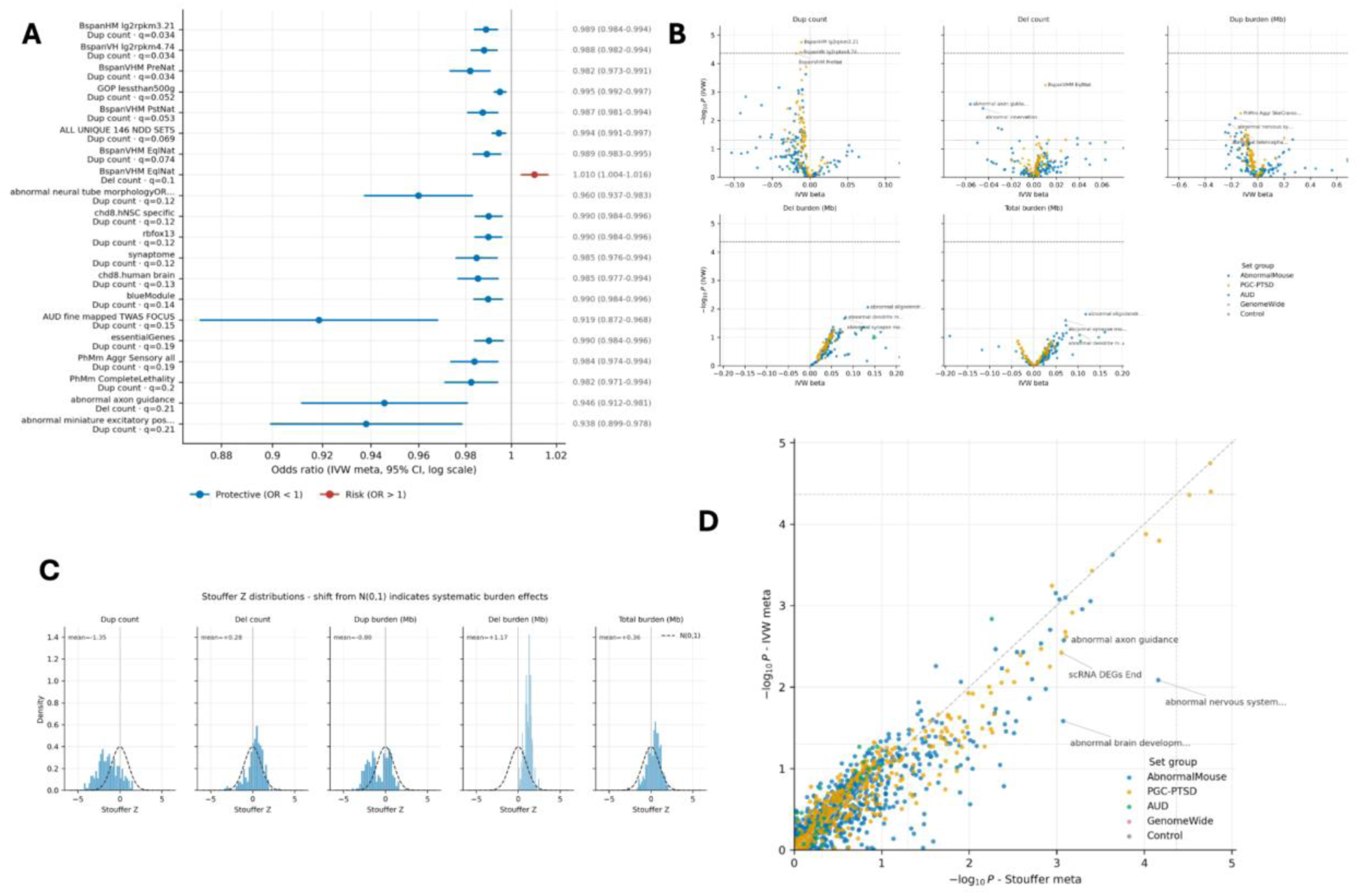
Cross-ancestry meta-analysis of gene-set rCNV burden in AUD (rCNVs of all sizes). **(A)** Odds ratios and 95% confidence intervals (log scale) for the 20 gene set–burden metric pairs with the smallest inverse-variance-weighted (IVW) meta-analysis *P*-values across European, African, and admixed American ancestry groups. Row labels give the gene set, burden metric, and FDR *q*-value; estimates are listed at right. Blue indicates odds ratios below 1 and red indicates odds ratios above 1. **(B)** Volcano plots of IVW effect size (beta) against −log10(*P*) for each burden metric: duplication count, deletion count, and duplication, deletion, and total burden (Mb). Points are colored by gene-set group. The dashed line marks the Bonferroni threshold and the dotted line marks *P* = 0.05; the most significant sets are labeled. **(C)** Distribution of Stouffer meta-analysis Z-scores across gene sets for each burden metric, compared with the standard normal distribution (dashed curve); the mean Z-score is shown in each panel. **(D)** Comparison of −log10(*P*) from Stouffer and IVW meta-analyses across all gene set–burden metric tests, colored by gene-set group. The diagonal indicates equal *P*-values; dashed and dotted lines mark the Bonferroni and nominal (*P* = 0.05) thresholds. All models were adjusted for ten principal components, age, sex, total rCNV count, and average rCNV length.

**Supplementary Figure 8.**
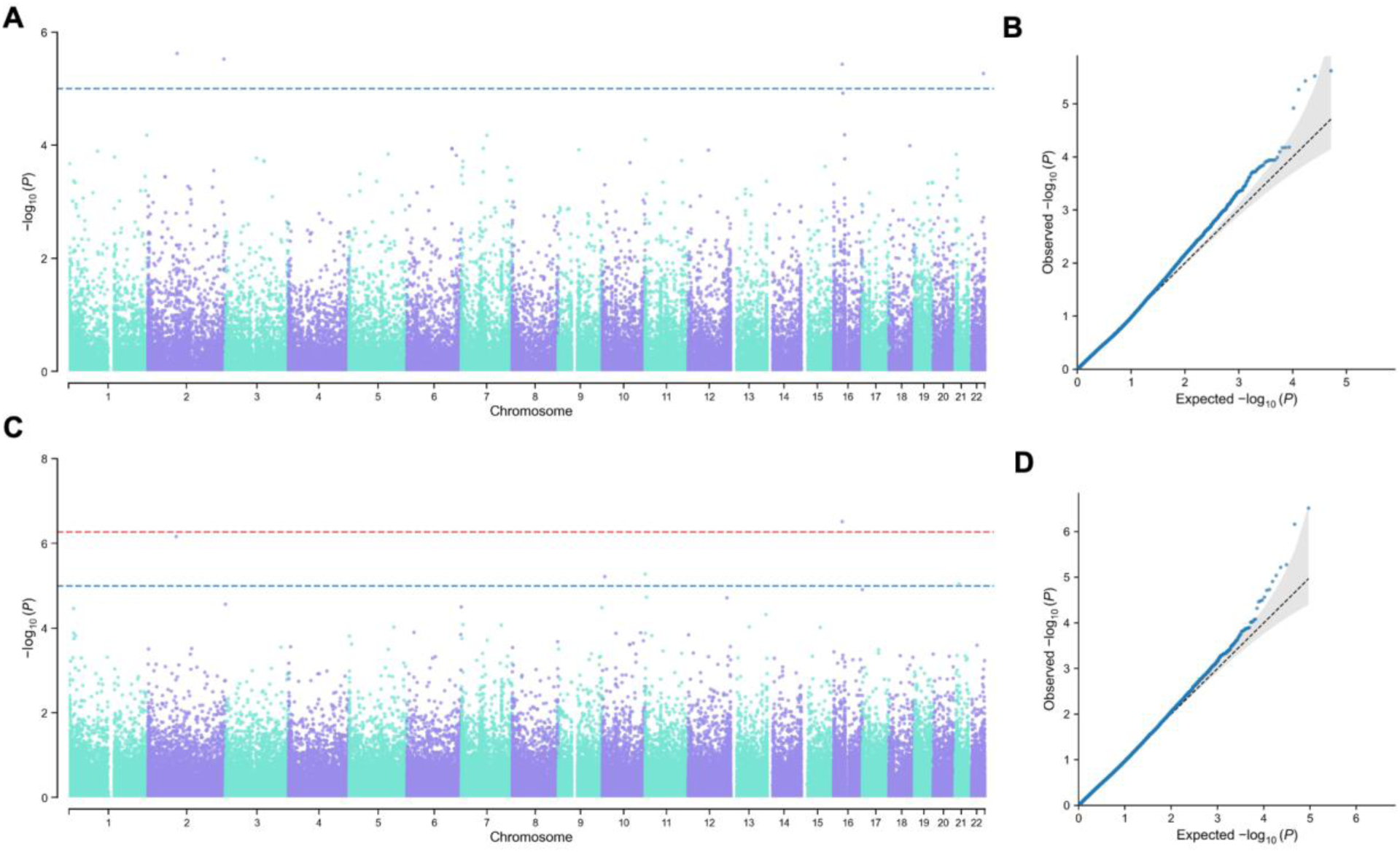
Region-level rCNV association with AUD. **(A, C)** Manhattan plots of −log10(*P*) by genomic position for the European ancestry (A) and trans-ancestry (C) analyses. The blue dashed line marks the suggestive threshold (*P* = 1 × 10⁻⁵) and the red dashed line marks the genome-wide significance threshold (*P* = 5.4 × 10⁻⁷). **(B, D)** Corresponding quantile–quantile plots of observed versus expected −log10(*P*) for the European ancestry (λ = 1.05) (B) and trans-ancestry (λ = 1.03) (D) analyses; the dashed line indicates the null expectation and the shaded area its 95% confidence interval.

## References

1. First MB. Diagnostic and statistical manual of mental disorders, 5th edition, and clinical utility. J Nerv Ment Dis. 2013 Sep;201(9):727–9. doi:10.1097/NMD.0b013e3182a2168a PubMed PMID: 23995026.

2. GBD 2016 Alcohol and Drug Use Collaborators. The global burden of disease attributable to alcohol and drug use in 195 countries and territories, 1990-2016: a systematic analysis for the Global Burden of Disease Study 2016. Lancet Psychiatry. 2018 Dec;5(12):987–1012. doi:10.1016/S2215-0366(18)30337-7 PubMed PMID: 30392731; PubMed Central PMCID: PMC6251968.

3. Haber PS. Identification and Treatment of Alcohol Use Disorder. N Engl J Med. 2025 Jan 16;392(3):258–66. doi:10.1056/NEJMra2306511 PubMed PMID: 39813644.

4. Choi HY, Balter DR, Haque LY. Epidemiology and Health Care Burden of Alcohol Use Disorder. Clin Liver Dis. 2024 Nov;28(4):577–88. doi:10.1016/j.cld.2024.06.006 PubMed PMID: 39362708; PubMed Central PMCID: PMC11772009.

5. Kranzler HR. Overview of Alcohol Use Disorder. AJP. 2023 Aug 1;180(8):565–72. doi:10.1176/appi.ajp.20230488

6. Verhulst B, Neale MC, Kendler KS. The heritability of alcohol use disorders: a meta-analysis of twin and adoption studies. Psychol Med. 2015 Apr;45(5):1061–72. doi:10.1017/S0033291714002165 PubMed PMID: 25171596; PubMed Central PMCID: PMC4345133.

7. 23andMe Research Team, Walters RK, Polimanti R, Johnson EC, McClintick JN, Adams MJ, et al. Transancestral GWAS of alcohol dependence reveals common genetic underpinnings with psychiatric disorders. Nat Neurosci. 2018 Dec;21(12):1656–69. doi:10.1038/s41593-018-0275-1

8. Kranzler HR, Zhou H, Kember RL, Vickers Smith R, Justice AC, Damrauer S, et al. Genome-wide association study of alcohol consumption and use disorder in 274,424 individuals from multiple populations. Nat Commun. 2019 Apr 2;10(1):1499. doi:10.1038/s41467-019-09480-8 PubMed PMID: 30940813; PubMed Central PMCID: PMC6445072.

9. Sanchez-Roige S, Palmer AA, Fontanillas P, Elson SL, 23andMe Research Team, the Substance Use Disorder Working Group of the Psychiatric Genomics Consortium, Adams MJ, et al. Genome-Wide Association Study Meta-Analysis of the Alcohol Use Disorders Identification Test (AUDIT) in Two Population-Based Cohorts. Am J Psychiatry. 2019 Feb 1;176(2):107–18. doi:10.1176/appi.ajp.2018.18040369 PubMed PMID: 30336701; PubMed Central PMCID: PMC6365681.

10. Saunders GRB, Wang X, Chen F, Jang SK, Liu M, Wang C, et al. Genetic diversity fuels gene discovery for tobacco and alcohol use. Nature. 2022 Dec;612(7941):720–4. doi:10.1038/s41586-022-05477-4 PubMed PMID: 36477530; PubMed Central PMCID: PMC9771818.

11. Zhou H, Kember RL, Deak JD, Xu H, Toikumo S, Yuan K, et al. Multi-ancestry study of the genetics of problematic alcohol use in over 1 million individuals. Nat Med. 2023 Dec;29(12):3184–92. doi:10.1038/s41591-023-02653-5 PubMed PMID: 38062264; PubMed Central PMCID: PMC10719093.

12. Zhou H, Sealock JM, Sanchez-Roige S, Clarke TK, Levey DF, Cheng Z, et al. Genome-wide meta-analysis of problematic alcohol use in 435,563 individuals yields insights into biology and relationships with other traits. Nat Neurosci. 2020 Jul;23(7):809–18. doi:10.1038/s41593-020-0643-5 PubMed PMID: 32451486; PubMed Central PMCID: PMC7485556.

13. Visscher PM, Wray NR, Zhang Q, Sklar P, McCarthy MI, Brown MA, et al. 10 Years of GWAS Discovery: Biology, Function, and Translation. Am J Hum Genet. 2017 Jul 6;101(1):5–22. doi:10.1016/j.ajhg.2017.06.005 PubMed PMID: 28686856; PubMed Central PMCID: PMC5501872.

14. Kang J, Deng YT, Wu BS, Liu WS, Li ZY, Xiang S, et al. Whole exome sequencing analysis identifies genes for alcohol consumption. Nat Commun. 2024 Jul 10;15(1):5777. doi:10.1038/s41467-024-50132-3 PubMed PMID: 38982111; PubMed Central PMCID: PMC11233704.

15. Ahangari M, Gentry AE, Hassan MF, Nguyen TH, Kendler KS, Bacanu SA, et al. Improving the discovery of rare variants associated with alcohol problems by leveraging machine learning phenotype prediction and functional information. bioRxiv. 2023 Sep 15;2023.09.11.557163. doi:10.1101/2023.09.11.557163 PubMed PMID: 37745400; PubMed Central PMCID: PMC10515858.

16. Hill SY, Hostyk J. A whole exome sequencing study to identify rare variants in multiplex families with alcohol use disorder. Front Psychiatry. 2023;14:1216493. doi:10.3389/fpsyt.2023.1216493 PubMed PMID: 37915799; PubMed Central PMCID: PMC10616827.

17. Rodríguez-López J, Flórez G, Blanco V, Pereiro C, Fernández JM, Fariñas E, et al. Genome wide analysis of rare copy number variations in alcohol abuse or dependence. J Psychiatr Res. 2018 Aug;103:212–8. doi:10.1016/j.jpsychires.2018.06.001 PubMed PMID: 29890507.

18. Ehlers CL, Wilhelmsen KC. Genomic scan for alcohol craving in Mission Indians. Psychiatr Genet. 2005 Mar;15(1):71–5. doi:10.1097/00041444-200503000-00012 PubMed PMID: 15722961.

19. Sulovari A, Liu Z, Zhu Z, Li D. Genome-wide meta-analysis of copy number variations with alcohol dependence. Pharmacogenomics J. 2018 May 22;18(3):398–405. doi:10.1038/tpj.2017.35 PubMed PMID: 28696413.

20. Hastings PJ, Lupski JR, Rosenberg SM, Ira G. Mechanisms of change in gene copy number. Nat Rev Genet. 2009 Aug;10(8):551–64. doi:10.1038/nrg2593 PubMed PMID: 19597530; PubMed Central PMCID: PMC2864001.

21. Molloy CJ, Quigley C, McNicholas Á, Lisanti L, Gallagher L. A review of the cognitive impact of neurodevelopmental and neuropsychiatric associated copy number variants. Transl Psychiatry. 2023 Apr 8;13(1):116. doi:10.1038/s41398-023-02421-6

22. Bacchelli E, Cameli C, Viggiano M, Igliozzi R, Mancini A, Tancredi R, et al. An integrated analysis of rare CNV and exome variation in Autism Spectrum Disorder using the Infinium PsychArray. Sci Rep. 2020 Feb 21;10(1):3198. doi:10.1038/s41598-020-59922-3 PubMed PMID: 32081867; PubMed Central PMCID: PMC7035424.

23. Viggiano M, Ceroni F, Visconti P, Posar A, Scaduto MC, Sandoni L, et al. Genomic analysis of 116 autism families strengthens known risk genes and highlights promising candidates. npj Genom Med. 2024 Mar 22;9(1):21. doi:10.1038/s41525-024-00411-1

24. Marshall CR, Howrigan DP, Merico D, Thiruvahindrapuram B, Wu W, Greer DS, et al. Contribution of copy number variants to schizophrenia from a genome-wide study of 41,321 subjects. Nat Genet. 2017 Jan;49(1):27–35. doi:10.1038/ng.3725

25. Montanucci L, Lewis-Smith D, Collins RL, Niestroj LM, Parthasarathy S, Xian J, et al. Genome-wide identification and phenotypic characterization of seizure-associated copy number variations in 741,075 individuals. Nat Commun. 2023 Jul 20;14(1):4392. doi:10.1038/s41467-023-39539-6

26. Kim J, Lee J, Jang DH. Combining chromosomal microarray and clinical exome sequencing for genetic diagnosis of intellectual disability. Sci Rep. 2023 Dec 20;13(1):22807. doi:10.1038/s41598-023-50285-z

27. Sønderby IE, Ching CRK, Thomopoulos SI, van der Meer D, Sun D, Villalon-Reina JE, et al. Effects of copy number variations on brain structure and risk for psychiatric illness: Large-scale studies from the ENIGMA working groups on CNVs. Hum Brain Mapp. 2022 Jan;43(1):300–28. doi:10.1002/hbm.25354 PubMed PMID: 33615640; PubMed Central PMCID: PMC8675420.

28. Cai N, Choi KW, Fried EI. Reviewing the genetics of heterogeneity in depression: operationalizations, manifestations and etiologies. Human Molecular Genetics. 2020 Sep 30;29(R1):R10–8. doi:10.1093/hmg/ddaa115

29. Deak JD, Levey DF, Wendt FR, Zhou H, Galimberti M, Kranzler HR, et al. Genome-Wide Investigation of Maximum Habitual Alcohol Intake in US Veterans in Relation to Alcohol Consumption Traits and Alcohol Use Disorder. JAMA Netw Open. 2022 Oct 3;5(10):e2238880. doi:10.1001/jamanetworkopen.2022.38880 PubMed PMID: 36301540; PubMed Central PMCID: PMC9614582.

30. Mallard TT, Savage JE, Johnson EC, Huang Y, Edwards AC, Hottenga JJ, et al. Item-Level Genome-Wide Association Study of the Alcohol Use Disorders Identification Test in Three Population-Based Cohorts. Am J Psychiatry. 2022 Jan;179(1):58–70. doi:10.1176/appi.ajp.2020.20091390 PubMed PMID: 33985350; PubMed Central PMCID: PMC9272895.

31. Mitchell AJ, Meader N, Bird V, Rizzo M. Clinical recognition and recording of alcohol disorders by clinicians in primary and secondary care: meta-analysis. Br J Psychiatry. 2012 Aug;201(2):93–100. doi:10.1192/bjp.bp.110.091199

32. Pocklington AJ, Rees E, Walters JTR, Han J, Kavanagh DH, Chambert KD, et al. Novel Findings from CNVs Implicate Inhibitory and Excitatory Signaling Complexes in Schizophrenia. Neuron. 2015 Jun 3;86(5):1203–14. doi:10.1016/j.neuron.2015.04.022 PubMed PMID: 26050040; PubMed Central PMCID: PMC4460187.

33. Nguyen HT, Bryois J, Kim A, Dobbyn A, Huckins LM, Munoz-Manchado AB, et al. Integrated Bayesian analysis of rare exonic variants to identify risk genes for schizophrenia and neurodevelopmental disorders. Genome Med. 2017 Dec 20;9(1):114. doi:10.1186/s13073-017-0497-y PubMed PMID: 29262854; PubMed Central PMCID: PMC5738153.

34. Pardiñas AF, Holmans P, Pocklington AJ, Escott-Price V, Ripke S, Carrera N, et al. Common schizophrenia alleles are enriched in mutation-intolerant genes and in regions under strong background selection. Nat Genet. 2018 Mar;50(3):381–9. doi:10.1038/s41588-018-0059-2 PubMed PMID: 29483656; PubMed Central PMCID: PMC5918692.

35. Maihofer AX, Engchuan W, Huguet G, Klein M, MacDonald JR, Shanta O, et al. Rare copy number variation in posttraumatic stress disorder. Mol Psychiatry. 2022 Dec;27(12):5062–9. doi:10.1038/s41380-022-01776-4

36. Singh M, Chatzinakos C, Barr PB, Gentry AE, Bigdeli TB, Webb BT, et al. Trans-ancestry Genome-Wide Analyses in UK Biobank Yield Novel Risk Loci for Major Depression. medRxiv. 2025 Feb 24;2025.02.22.25322721. doi:10.1101/2025.02.22.25322721 PubMed PMID: 40061314; PubMed Central PMCID: PMC11888526.

37. Mann K, Lehert P, Morgan MY. The Efficacy of Acamprosate in the Maintenance of Abstinence in Alcohol-Dependent Individuals: Results of a Meta-Analysis. Alcoholism Clin & Exp Res. 2004 Jan;28(1):51–63. doi:10.1097/01.ALC.0000108656.81563.05

38. Skinner MD, Lahmek P, Pham H, Aubin HJ. Disulfiram Efficacy in the Treatment of Alcohol Dependence: A Meta-Analysis. Laks J, editor. PLoS ONE. 2014 Feb 10;9(2):e87366. doi:10.1371/journal.pone.0087366

39. Sudakin D. Naltrexone: Not Just for Opioids Anymore. J Med Toxicol. 2016 Mar;12(1):71–5. doi:10.1007/s13181-015-0512-x

40. Peterson RE, Edwards AC, Bacanu S, Dick DM, Kendler KS, Webb BT. The utility of empirically assigning ancestry groups in cross-population genetic studies of addiction. American J Addict. 2017 Aug;26(5):494–501. doi:10.1111/ajad.12586

41. Koenig Z, Yohannes MT, Nkambule LL, Zhao X, Goodrich JK, Kim HA, et al. A harmonized public resource of deeply sequenced diverse human genomes. Genome Res. 2024 May;34(5):796–809. doi:10.1101/gr.278378.123

42. Raychaudhuri S, Korn JM, McCarroll SA, International Schizophrenia Consortium, Altshuler D, Sklar P, et al. Accurately assessing the risk of schizophrenia conferred by rare copy-number variation affecting genes with brain function. PLoS Genet. 2010 Sep 9;6(9):e1001097. doi:10.1371/journal.pgen.1001097 PubMed PMID: 20838587; PubMed Central PMCID: PMC2936523.

43. Frankish A, Carbonell-Sala S, Diekhans M, Jungreis I, Loveland JE, Mudge JM, et al. GENCODE: reference annotation for the human and mouse genomes in 2023. Nucleic Acids Res. 2023 Jan 6;51(D1):D942–9. doi:10.1093/nar/gkac1071 PubMed PMID: 36420896; PubMed Central PMCID: PMC9825462.

44. Demerath NJ. THE AMERICAN SOLDIER: VOLUME I, ADJUSTMENT DURING ARMY LIFE. By S. A. Stouffer, E. A. Suchman, L. C. DeVinney, S. A. Star, R. M. Williams, Jr. VOLUME II, COMBAT AND ITS AFTERMATH. By S. A. Stouffer, A. A. Lumsdaine, M. H. Lumsdaine, R. M. Williams, Jr., M. B. Smith, I. L. Janis, S. A. Star, L. S. Cottrell, Jr. Princeton, New Jersey: Princeton University Press, 1949. Vol. I, 599 pp., Vol. II, 675 pp. $7.50 each; $13.50 together. Social Forces. 1949 Oct 1;28(1):87–90. doi:10.2307/2572105

45. Willer CJ, Li Y, Abecasis GR. METAL: fast and efficient meta-analysis of genomewide association scans. Bioinformatics. 2010 Sep 1;26(17):2190–1. doi:10.1093/bioinformatics/btq340

46. Girirajan S, Rosenfeld JA, Cooper GM, Antonacci F, Siswara P, Itsara A, et al. A recurrent 16p12.1 microdeletion supports a two-hit model for severe developmental delay. Nat Genet. 2010 Mar;42(3):203–9. doi:10.1038/ng.534

47. Jensen M, Smolen C, Tyryshkina A, Pizzo L, Sun J, Noss S, et al. Genetic modifiers and ascertainment drive variable expressivity of complex disorders. Cell. 2025 Dec 11;188(25):7065–7082.e17. doi:10.1016/j.cell.2025.09.012 PubMed PMID: 41061703; PubMed Central PMCID: PMC12614355.

48. Smolen C, Jensen M, Dyer L, Pizzo L, Tyryshkina A, Banerjee D, et al. Assortative mating and parental genetic relatedness drive the pathogenicity of variably expressive variants [Internet]. Genetic and Genomic Medicine; 2023 [cited 2026 Jul 29]. Available from: http://medrxiv.org/lookup/doi/10.1101/2023.05.18.23290169 doi:10.1101/2023.05.18.23290169

49. Wang L, Kranzler HR, Gelernter J, Zhou H. Investigating the Contribution of Coding Variants in Alcohol Use Disorder Using Whole-Exome Sequencing Across Ancestries. Biological Psychiatry. 2025 Jul;98(1):46–55. doi:10.1016/j.biopsych.2025.01.020

50. Hindle SJ, Munji RN, Dolghih E, Gaskins G, Orng S, Ishimoto H, et al. Evolutionarily Conserved Roles for Blood-Brain Barrier Xenobiotic Transporters in Endogenous Steroid Partitioning and Behavior. Cell Rep. 2017 Oct 31;21(5):1304–16. doi:10.1016/j.celrep.2017.10.026 PubMed PMID: 29091768; PubMed Central PMCID: PMC5774027.

51. Kirov G, Rees E, Walters JTR, Escott-Price V, Georgieva L, Richards AL, et al. The penetrance of copy number variations for schizophrenia and developmental delay. Biol Psychiatry. 2014 Mar 1;75(5):378–85. doi:10.1016/j.biopsych.2013.07.022 PubMed PMID: 23992924; PubMed Central PMCID: PMC4229045.

52. Jain L, Oberman LM, Beamer L, Cascio L, May M, Srikanth S, et al. Genetic and metabolic profiling of individuals with Phelan-McDermid syndrome presenting with seizures. Clin Genet. 2022 Jan;101(1):87–100. doi:10.1111/cge.14074 PubMed PMID: 34664257.

53. Sugawara H, Murata Y, Ikegame T, Sawamura R, Shimanaga S, Takeoka Y, et al. DNA methylation analyses of the candidate genes identified by a methylome-wide association study revealed common epigenetic alterations in schizophrenia and bipolar disorder. Psychiatry Clin Neurosci. 2018 Apr;72(4):245–54. doi:10.1111/pcn.12645 PubMed PMID: 29430824.

54. Mehraein Y, Pfob M, Steinlein O, Aichinger E, Eggert M, Bubendorff V, et al. 2q37.3 Deletion Syndrome: Two Cases with Highly Distinctive Facial Phenotype, Discordant Association with Schizophrenic Psychosis, and Shared Deletion Breakpoint Region on 2q37.3. Cytogenet Genome Res. 2015;146(1):33–8. doi:10.1159/000431389 PubMed PMID: 26112830.

55. Ogura K, Takeshita K, Arakawa C, Shimojima K, Yamamoto T. Neuropsychological profiles of patients with 2q37.3 deletion associated with developmental dyspraxia. Am J Med Genet B Neuropsychiatr Genet. 2014 Dec;165B(8):684–90. doi:10.1002/ajmg.b.32274 PubMed PMID: 25329715.

